# BCG vaccination recalibrates innate immunity in ART-treated people with HIV

**DOI:** 10.64898/2026.08.28.26361620

**Authors:** Cédric Dollé, Terence K. Tutumlu, Lena Bartl, Baptiste Depouilly, Doris Russenberger, Marius Zeeb, Katharina Kusejko, Emily West, Dominique L. Braun, Magdalena Schwarzmüller, Baptiste Elie, Alexandra Trkola, Huldrych F. Günthard, Johannes Nemeth

## Abstract

Despite suppressive antiretroviral therapy, many people with HIV (PWH) retain chronic interferon-associated immune dysregulation. Observational data from the Swiss HIV Cohort Study linked asymptomatic mycobacterial exposure to lower viral set points, reduced interferon-associated activity, and attenuated HIV-specific antibody responses, a pattern sharing features with HIV elite controllers and natural hosts of primate lentiviruses. We therefore examined whether Bacillus Calmette-Guérin (BCG) vaccination could induce a related immune configuration in ART-treated PWH. Using longitudinal systems-level profiling within the BELIEVE trial, we found that BCG reduced constitutive NK cell IFN-γ production and PBMC-mediated direct cytotoxicity without impairing inducible cytokine responses or antibody-dependent cellular cytotoxicity. Multiomic and proteomic analyses showed reduced interferon- and activation-associated programs, while adaptive immune parameters remained largely stable and follow-up revealed no obvious adverse clinical pattern. This configuration, reduced baseline interferon activity coexisting with preserved Fc-dependent effector function, shares selected features with immune states described in natural lentiviral control and provides a rationale for testing BCG in combination with antibody-based HIV interventions.

## Introduction

Despite effective viral suppression, antiretroviral therapy (ART) does not fully restore immune homeostasis in many people with HIV (PWH). Chronic immune activation and persistent interferon signaling can remain detectable during suppressive ART [1–4]. NK cells are of particular interest in this setting, as HIV infection has been associated with durable alterations in NK cell phenotype, activation state, and cytotoxic potential, some of which persist despite ART [4–6]. This raises the question of whether chronic immune activation can be reduced without compromising antiviral effector function.

Natural lentiviral infection illustrate that long-term viral persistence or control can occur without progressive disease. In African green monkeys and sooty mangabeys, SIV infection remains non-pathogenic despite persistent high-level viremia and is accompanied by rapid resolution of the acute type I interferon response, while NK cells in African green monkeys accumulate in lymph-node follicles and contribute to local control of viral replication [7–9]. In humans, elite controllers provide a distinct example of durable HIV control without ART. In stringently defined controller cohorts, this control has been associated with comparatively low systemic inflammation alongside strong HIV-specific cytotoxic responses, although systemic interferon and innate activation profiles remain heterogeneous across studies [10–13]. Together, these observations suggest that lentiviral control or tolerance can coexist with preserved antiviral function without requiring uniformly high systemic immune activation. Whether aspects of this balance can be deliberately induced remains an open question.

Observational studies from the Swiss HIV Cohort Study (SHCS) further suggest that asymptomatic *Mycobacterium tuberculosis* (Mtb) infection can alter the immune environment in PWH. Mtb infection was associated with lower HIV-1 viral set points and a lower risk of opportunistic infections, while transcriptomic profiling identified reduced IFN-α- and IFN-γ-associated pathway activity alongside selective increases in other inflammatory programs [14,15]. Mtb infection was also associated with reduced HIV-specific antibody responses [16]. Although these studies do not establish causality, they indicate that mycobacterial exposure can be accompanied by coordinated changes in innate and adaptive immunity relevant to HIV infection. Bacillus Calmette-Guérin (BCG) vaccination provides a controlled setting in which to examine whether mycobacterial exposure can similarly reshape systemic immunity in ART-treated PWH.

Beyond protection against tuberculosis, BCG exerts heterologous immunological effects, an observation already proposed by Calmette in 1931 [17]. These effects are commonly discussed within the framework of trained immunity, involving sustained changes in cytokine responsiveness, epigenetic regulation, and cellular metabolism [18,19]. In healthy volunteers, BCG has been shown to enhance heterologous monocyte responses, modify NK cell function, and alter hematopoietic stem and progenitor cells [20–22]. However, more recent studies indicate that BCG responses are heterogeneous and can also involve reduced steady-state inflammation with preserved or reshaped responses to secondary challenge, depending partly on the pre-existing immune state of the host [23–25]. BCG may therefore reorganize immune function rather than uniformly amplify innate activation.

The immune landscape of treated HIV differs from that of the immunologically healthy populations in which most BCG studies have been performed, raising the possibility that vaccination may produce a distinct response in ART-treated PWH. BCG has not been extensively studied in PWH, partly because of historical safety concerns surrounding live mycobacterial vaccination in immunocompromised hosts [26]. The BELIEVE trial addressed this knowledge gap by demonstrating the clinical safety of BCG in virologically suppressed PWH with immune reconstitution and providing a longitudinal framework to examine its broader immunological effects [27].

Here, using longitudinal systems-level immune profiling embedded within the BELIEVE trial, we examined how BCG vaccination reshapes systemic immune function in ART-treated PWH across innate and adaptive compartments.

## Results

### Myeloid cell proportions increase following BCG vaccination

Longitudinal samples from 55 BELIEVE participants [27] were assessed at baseline (day 0) and at 90 and 180 days after BCG vaccination (Fig. 1a). To determine whether BCG vaccination altered the composition of the circulating immune compartment, PBMCs were analyzed using high-dimensional flow cytometry (Supplementary Table 1) followed by automated clustering and manual annotation. This approach identified five major cell populations (Fig. 1b, SF1a), with proportions largely within expected ranges. While lymphocyte proportions remained stable over time, myeloid cell proportions increased at both post-vaccination timepoints despite substantial inter-individual variability (Fig. 1c-d). This pattern was corroborated by absolute cell counts, which showed a concordant increase in myeloid cells (SF1b).

**Figure 1.**
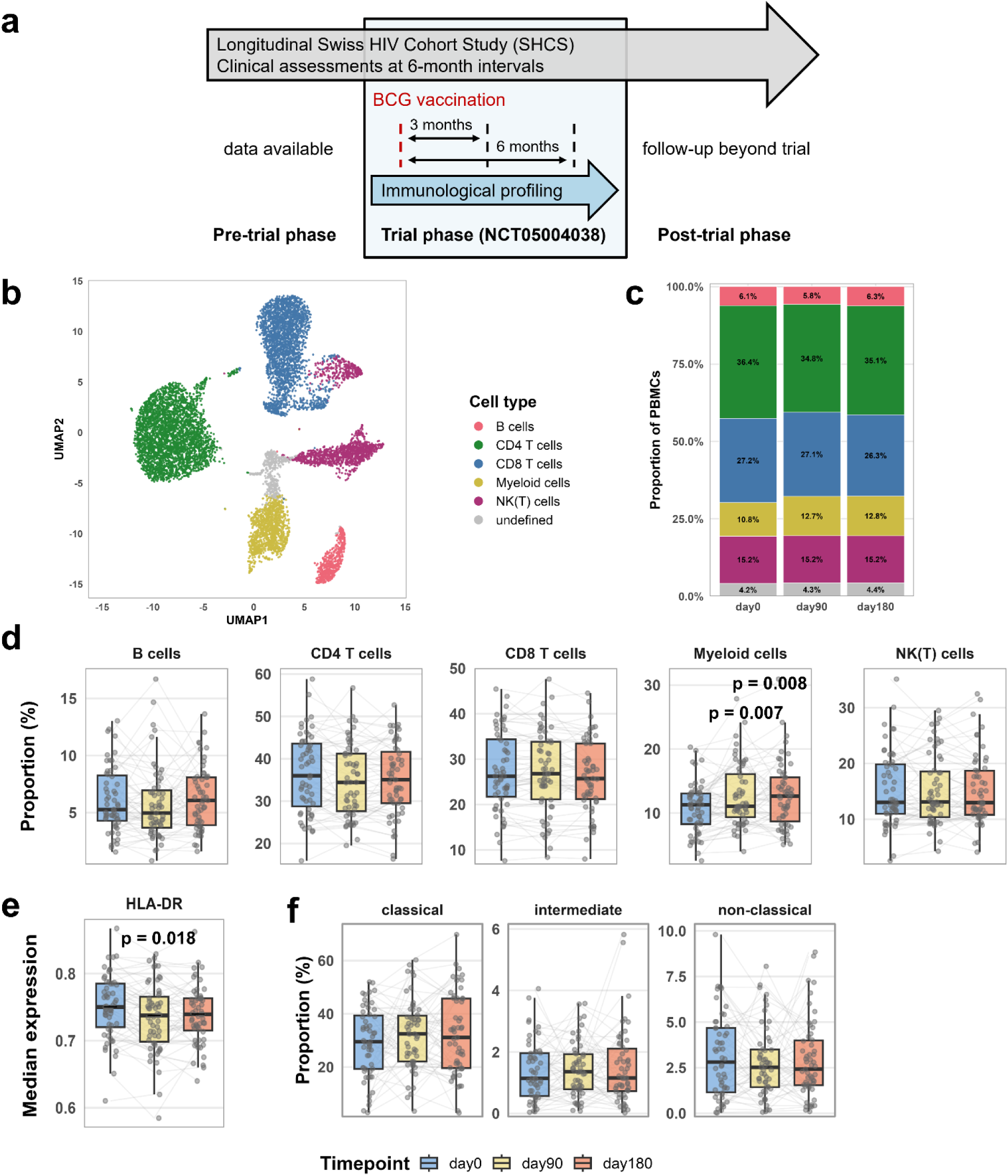
Increase in circulating myeloid-cell proportions after BCG vaccination. a) Schematic overview of the clinical trial (NCT05004038) within the Swiss HIV Cohort Study (SHCS). b) UMAP showing immune cell populations identified from PBMCs by flow cytometry-based clustering and annotation. c) Stacked bar plots showing proportions of immune cell populations across all three study timepoints. d) Boxplots showing cell type proportions across timepoints, with lines and points representing individual participants (n = 55). e) Boxplots showing median HLA-DR expression in CD33+ myeloid cells across timepoints, with lines and points representing individual participants (n = 55). f) Boxplots showing monocyte subtype proportions across timepoints, with lines and points representing individual participants (n = 55). Statistics: Differential abundance in panels d and f was assessed using diffcyt-DA-GLMM, and differential state in panel e using diffcyt-DS-LMM. P values were FDR-adjusted; only adjusted p < 0.05 are indicated. Data were derived from flow cytometry.

To further assess functional phenotypes, we examined median expression of markers not used for clustering. Among these, only HLA-DR showed a consistent decrease in myeloid cells (Fig. 1e), whereas marker expression in other major cell types remained unchanged (SF2-3). These findings indicate that although myeloid cells increase in abundance following BCG vaccination, they do not exhibit signs of enhanced activation.

To assess the robustness of this observation, we repeated the clustering using a broader marker set (fully automated without manual annotation). Among 15 clusters, only one cluster (cluster 10) showed significant changes over time (SF4a-b). This cluster displayed a CD33^+^ HLA-DR^+^ CD38^+^ CD11c^dim^ phenotype consistent with antigen presenting myeloid cells, supporting the primary analysis.

Finally, subclustering of CD33^+^ myeloid cells based on CD14, CD16, and CD11c revealed no significant shifts in major monocyte subsets, despite a trend toward increased classical monocytes (Fig. 1f).

### No evidence of a trained immunity phenotype in myeloid cells

To determine whether BCG vaccination induces trained immunity in PWH, we assessed if myeloid cells exhibited enhanced TNF-α responses upon restimulation as a commonly used functional readout for trained immunity. PBMCs were stimulated for 4 h with LPS, β-glucan, or heat-killed *M. tuberculosis* (hk-Mtb), and the proportion of TNF-α^+^ myeloid cells and monocyte subsets was quantified across timepoints. TNF-α responses remained unchanged at day 90 and day 180 compared to baseline across all stimuli (SF5+SF6). This was consistent irrespective of whether responses were normalized to baseline or to the unstimulated condition at the respective timepoint. In line with the observed decrease in HLA-DR expression, these findings argue against a trained immunity-like phenotype in circulating myeloid cells.

### BCG vaccination differentially affects NK cell effector functions

To obtain an integrated view of immune co-variation across time, we summarized significant correlations between flow cytometry-derived features at the level of major immune compartments. At baseline, the meta-correlation network showed several inter-compartment connections (Fig. 2a). After BCG vaccination, the innate-cell connectivity appeared reduced, with fewer retained correlations involving the myeloid and NK(T) compartments at day 90 and day 180. Although this analysis is descriptive and does not infer directionality, it suggested a reorganization of immune co-variation after BCG vaccination, particularly involving innate immune compartments. This broader network-level pattern prompted closer examination of individual NK cell functional readouts.

**Figure 2.**
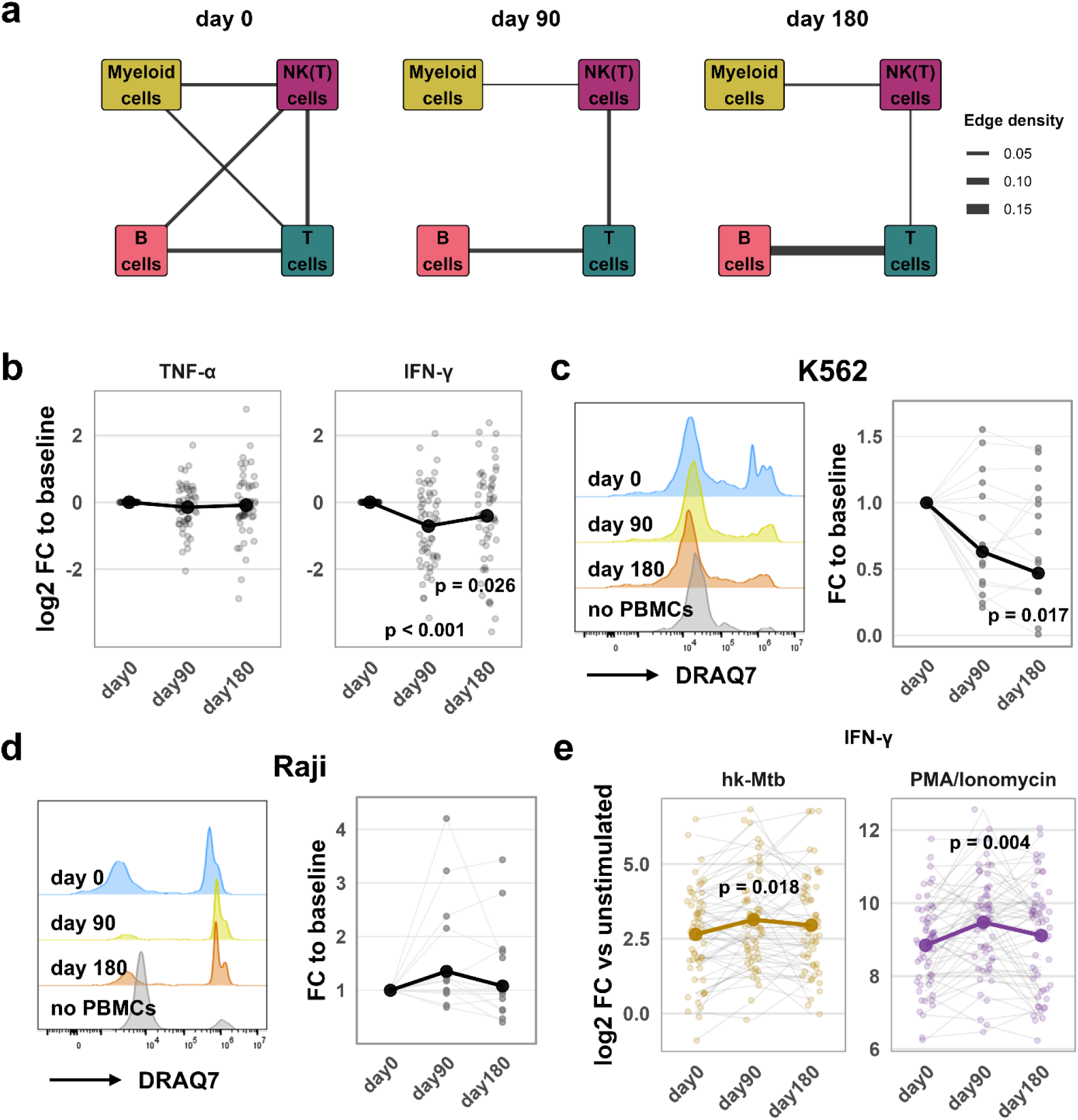
Differential effects of BCG vaccination on NK cell cytokine production, direct target-cell killing, and antibody-dependent cytotoxicity. a) Meta-correlation networks of flow cytometry-derived immune features at day 0, day 90, and day 180. Edges represent significant inter-compartment Spearman correlations between features (|r| > 0.4, FDR-adjusted p < 0.05), and edge width indicates the density of significant correlations among all possible feature pairs between two compartments. b) Log2 fold-changes relative to baseline (day 0) of TNF-α- and IFN-γ-producing CD56+ cells following 4 h brefeldin A treatment under unstimulated conditions. Lines represent estimated marginal means and points represent individual participants (n = 55). c) Representative flow cytometry histogram showing DRAQ7 staining used to quantify target-cell death after 4 h co-culture of K562 cells with PBMCs. Fold-changes relative to baseline are shown. Thick line represents estimated marginal means and points with connecting thin lines represent individual participants (n = 15). d) Representative flow cytometry histogram showing DRAQ7 staining used to quantify target-cell death after 6 h co-culture of Raji cells with PBMCs. Fold-changes relative to baseline are shown. Thick line represents estimated marginal means and points with connecting thin lines represent individual participants (n = 15). e) Log2 fold-changes relative to the corresponding unstimulated condition of TNF-α- and IFN-γ-producing CD56⁺ cells following 4 h brefeldin A treatment under heat-killed M. tuberculosis (hk-Mtb) and PMA/Ionomycin stimulation. Thick line represents estimated marginal means and points with connecting thin lines represent individual participants (n = 55). Statistics: Longitudinal changes were assessed using linear mixed-effects models, with planned contrasts of day 90 and day 180 vs. day 0. P values were FDR-adjusted; adjusted p < 0.05 are indicated. Data were derived from flow cytometry.

In contrast to the largely stable cytokine responses in myeloid cells and conventional adaptive lymphocytes, CD56^+^ NK(T) cells exhibited pronounced changes. Specifically, the proportion of IFN-γ^+^ cells under unstimulated conditions decreased by 39% at day 90 (log2FC −0.71) and remained reduced by 25% at day 180 (log2FC −0.41) (Fig. 2b), whereas both TNF-α and IFN-γ production remained largely unchanged across all other cell types (SF7). Further distinction of NK(T) cells using CD3 showed that the decrease was more pronounced in NK cells than in NKT cells (SF8a).

To determine whether reduced cytokine production was accompanied by altered effector function, we performed cytotoxicity assays using PBMCs co-cultured with K562 target cells. Consistent with the cytokine data, PBMC-mediated target cell killing showed reductions of 37% at day 90 and 53% at day 180 (log2FC −0.67 and −1.09, respectively) (Fig. 2c, SF8b). Depletion of CD56^+^ cells reduced target cell killing by an average of 80%, indicating that cytotoxic activity was predominantly mediated by CD56^+^ cells (SF8c). We then extended these analyses to additional target cell lines. Killing of ACH-2 cells, which harbor latent HIV-1, was reduced by 31-34%, while killing of their parental A3.01 cell line was reduced by 14-20% [28], indicating that the reduction in PBMC-mediated killing was not restricted to a single target model (SF8d-f). Overall, the direction of change was consistent across target cell lines, although the magnitude of the reduction varied.

To further characterize NK cells, we assessed the proportions of cells expressing granzyme B (GzmB), a cytolytic effector molecule, and CD16, an Fc receptor involved in antibody-mediated effector functions, as well as the distribution of CD56bright and CD56dim NK cell subsets, which are broadly associated with cytokine production and cytotoxic effector function, respectively [29]. None of these subsets showed major phenotypic shifts (SF9).

We next assessed rituximab-dependent killing of Raji cells in PBMC co-cultures. In contrast to the decrease in direct cytotoxicity, rituximab-dependent killing increased by 35% at day 90 compared with baseline (log2FC 0.43; p = 0.034) before returning toward baseline at day 180 (Fig. 2d, SF10a). This day-90 contrast did not remain significant after false discovery rate (FDR) correction (adjusted p = 0.067) across both post-vaccination timepoints, suggesting a possible transient increase in antibody-dependent cellular cytotoxicity (ADCC) after BCG vaccination. CD56 depletion reduced rituximab-dependent killing by an average of 39%, indicating a considerable but not exclusive contribution of CD56^+^ cells in this whole-PBMC assay (SF10b). Upon stimulation with hk-Mtb or PMA/Ionomycin, IFN-γ production remained inducible and comparable across timepoints (SF10c). When normalized to the corresponding unstimulated condition, IFN-γ responses were increased by 41% following hk-Mtb stimulation (log2FC 0.49) and by 55% following PMA/Ionomycin stimulation (log2FC 0.63) at day 90 (Fig. 2e), suggesting preserved inducible function and a relative shift toward stimulated responses despite reduced baseline activity.

### Single-cell multiome profiling of NK cells suggests reduced interferon signaling and shifts in transcriptional states

To explore whether the NK cell phenotype was accompanied by transcriptional or regulatory changes, we performed single-cell multiome profiling of CD56^+^ cells in an exploratory pooled analysis. To mirror the cytotoxicity assay conditions, PBMCs from three donors were co-cultured with K562 target cells. Baseline and day 90 samples were pooled separately, enriched for CD56^+^ cells, and then processed for paired gene expression (RNA) and chromatin accessibility (ATAC) analysis.

Gene expression analysis identified six Seurat clusters, with clusters 0-2 comprising more than 75% of all cells (Fig. 3A, SF11a). Mapping these clusters to previously described NK cell subsets [29] revealed comparable proportions of NK1-NK3 subsets between baseline and day 90, suggesting that the observed functional changes were not primarily explained by major shifts in canonical NK subset composition (SF11b). Instead, post-vaccination changes were most apparent at the level of transcriptionally defined states. Cluster 0 was characterized by cytokine-responsive genes including IL12RB2, KLRC1, ZBTB16, and XCL2, cluster 1 by immediate early activation genes including FOS, FOSB, JUN, and JUNB, and cluster 2 by interferon-stimulated genes including RSAD2, IFI44, IFI6, and OAS3 (SF11c). In the pooled day-90 sample, cluster 0 was more abundant and cluster 2 less abundant than in the pooled baseline sample, indicating a redistribution among NK cell transcriptional states (Fig. 3b).

**Figure 3.**
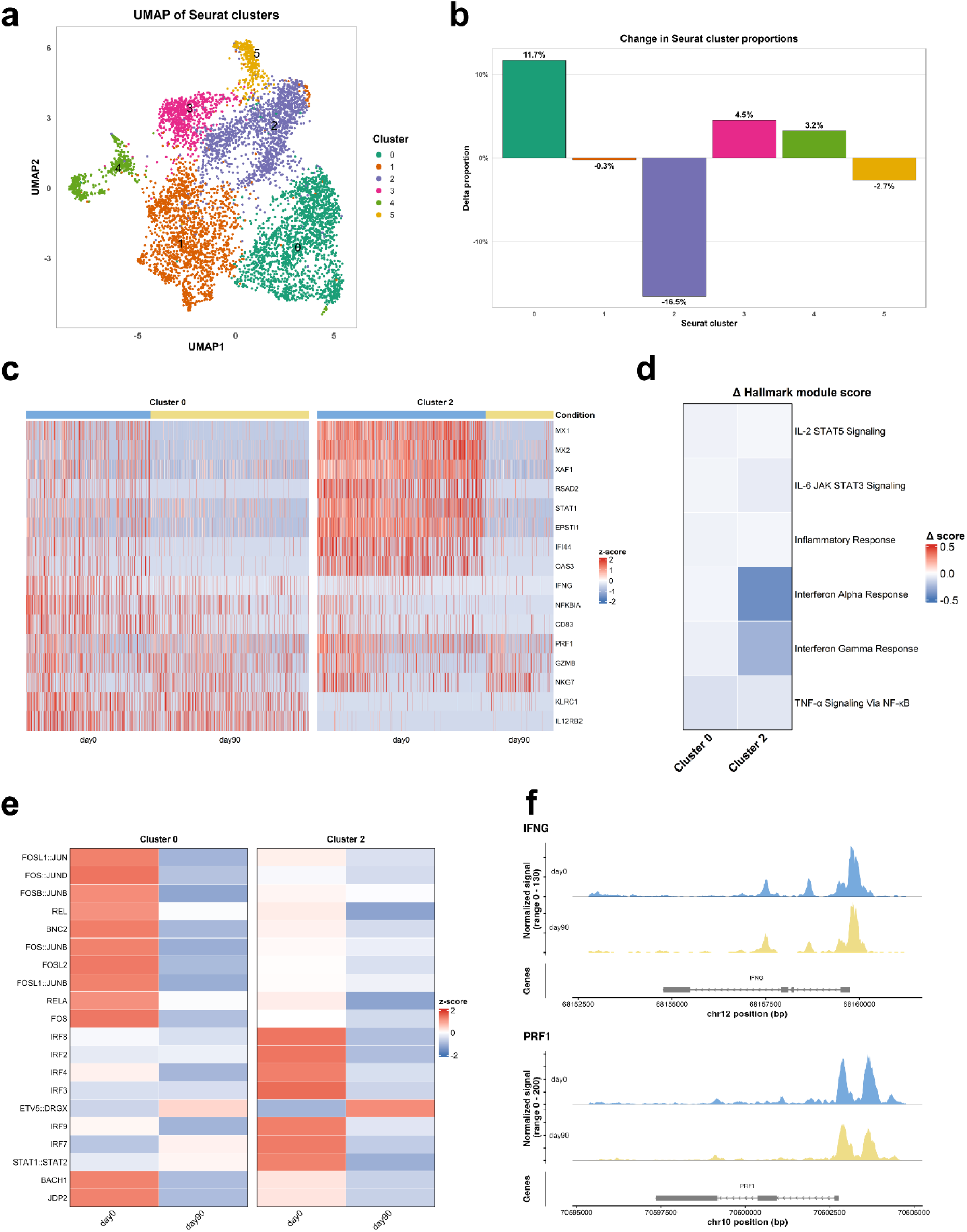
Single-cell multiome profiling of NK cell states before and after BCG vaccination. a) UMAP visualization of RNA-based Seurat clusters (0-5). b) Relative change in cluster proportions between day 0 and day 90, shown as percentage change for each cluster. Positive values indicate a higher proportion at day 90, whereas negative values indicate a lower proportion at day 90. c) Heatmap showing single-cell expression of selected genes in clusters 0 and 2 at day 0 and day 90. Genes were selected based on the strongest changes between timepoints together with canonical NK cell markers and cytokine-related genes. d) Heatmap of selected Hallmark gene sets showing changes in module scores between day 90 and day 0 (Δ score) in clusters 0 and 2. Module scores are shown as the difference in median score between post- and pre-vaccination cells within each cluster. e) Heatmap of top differential transcription factor motif activities in clusters 0 and 2, shown as z-scored median chromVAR deviation scores at day 0 and day 90. f) Chromatin accessibility coverage plots for the IFNG and PRF1 loci, showing aggregated ATAC signal at day 0 and day 90. Statistics: Samples from three donors were pooled per timepoint; therefore, no donor-level inferential testing was performed, and results are shown descriptively. Single-cell RNA and ATAC measurements were derived from single-cell multiome profiling.

Across clusters, transcriptional changes were dominated by reduced expression of interferon-stimulated genes, including MX1, MX2, RSAD2, IFI6, IFI44, and STAT1 (Fig. 3c, SF12a). Consistently, pathway-level analysis using curated immune gene sets showed lower type I and type II interferon response signatures at day 90 compared with baseline (Fig. 3d).

Chromatin accessibility analysis provided concordant regulatory support (SF12b). In cluster 2, motif activity for interferon-associated IRF and STAT family transcription factors, including IRF8, IRF7, and STAT1::STAT2, was reduced after vaccination (Fig. 3e). In cluster 0, which increased in relative abundance, AP-1 and NF-κB motif activity, including FOS::JUN, BATF::JUN, REL, and RELA, was lower (Fig. 3f). These findings are consistent with attenuation of interferon- and activation-linked regulatory programs. Accessibility at selected effector loci, including IFNG and PRF1, showed a similar direction of change (SF12c).

Together, these exploratory multiomic data are consistent with the functional observations and suggest that reduced NK cell IFN-γ production and PBMC-mediated cytotoxicity are accompanied by lower interferon- and activation-associated regulatory programs after BCG vaccination.

### Interferon-associated changes occur without broad alterations in adaptive immune function

To assess whether the NK cell and interferon-related changes observed at the cellular level were reflected more broadly, we analyzed systemic and adaptive immune responses.

Plasma proteomic profiling showed limited overall protein-level changes at 90 or 180 days following BCG vaccination. However, pathway-level analysis using immune-related gene sets identified negative enrichment of IFN-associated proteins, including IFN-γ and IFN-α responses, with the clearest and FDR-significant signal observed at day 90 (Fig. 4a). Examination of individual proteins within this set showed a general decrease over time (Fig. 4b), consistent with the reduced IFN-γ production and interferon-related signatures observed in NK cells.

**Figure 4.**
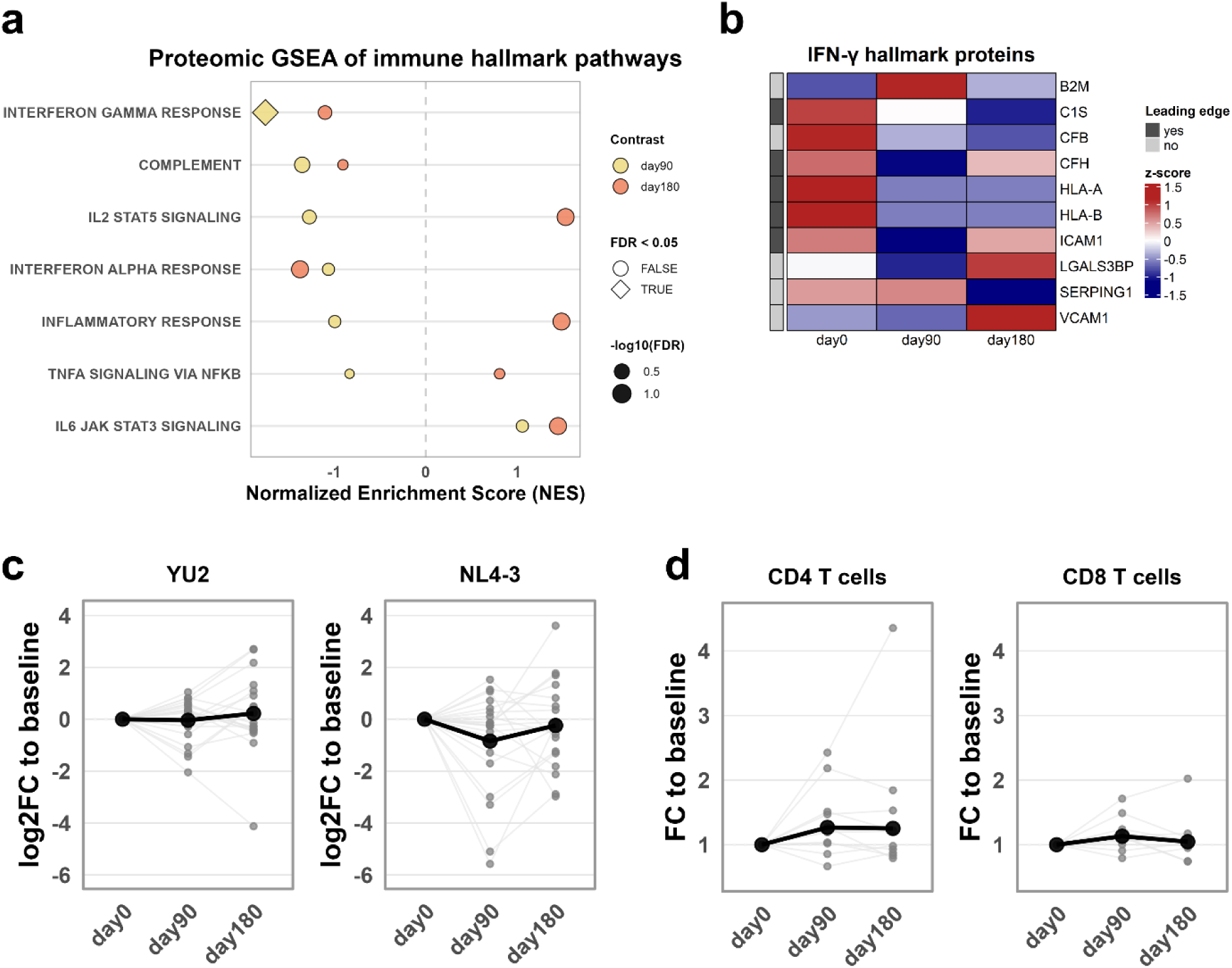
IFN-γ-related proteomic changes with stable T cell proliferation and PBMC susceptibility to HIV infection. a) Gene set enrichment analysis (GSEA) of immune-related Hallmark pathways using longitudinal proteomic data. Normalized enrichment scores (NES) are shown for day 90 vs. day 0 and day 180 vs. day 0. Symbol shape indicates significance (diamonds for FDR-adjusted p < 0.05; circles for FDR-adjusted p ≥ 0.05), and symbol size reflects −log10(FDR-adjusted p) (n = 55). b) Heatmap of detected proteins associated with the IFN-γ response Hallmark gene set. Shown are z-scored protein abundances across all three timepoints. Proteins contributing to the leading-edge subset in the GSEA are indicated. c) Log2 fold-changes relative to baseline (day 0) of PBMC susceptibility to HIV infection measured by TZM-bl luminescence following in vitro infection with HIV-1 strains YU2 and NL4-3 (MOI 0.01). Thick line represents estimated marginal means and points with connecting thin lines represent individual participants (n = 20). d) Fold-changes relative to baseline (day 0) of proliferating CD4+ and CD8+ T cells, identified by Tag-it Violet dilution after 96 h incubation with CD3/CD28 T cell activation beads. Thick line represents estimated marginal means and points with connecting thin lines represent individual participants (n = 10). Statistics: Pathway enrichment in panel a was assessed using GSEA with FDR correction across the predefined immune-related Hallmark pathways. Longitudinal changes in panels c and d were assessed using linear mixed-effects models, with planned contrasts of day 90 and day 180 vs. day 0. P values were FDR-adjusted; only adjusted p < 0.05 are indicated. Data were derived from mass spectrometry-based plasma proteomics (a, b), a TZM-bl reporter assay (c), and flow cytometry (d).

We then assessed whether this reduced activation state translated into altered susceptibility to viral infection. Activated PBMCs were infected with HIV-1 strains YU2 or NL4-3, and production of infectious virus was quantified using a TZM-bl reporter assay. Infection levels did not differ between timepoints for either viral strain (Fig. 4c, SF13), indicating that the observed immunological changes were not associated with increased permissiveness to HIV replication in this system.

To further evaluate adaptive immune function, we performed proliferation assays and stimulation experiments. Neither CD4^+^ nor CD8^+^ T cells showed significant changes in proliferation following stimulation with anti-CD3/CD28 beads (Fig. 4d). Cytokine responses following 4 h stimulation showed modest but consistent directional changes across selected conditions. In particular, reduced proportions of TNF-α-producing B cells following PMA/Ionomycin stimulation and IFN-γ-producing CD8 T cells following hk-Mtb stimulation were observed at post-vaccination timepoints (SF14+SF15). These changes were restricted to individual cell type-stimulus combinations and did not indicate a broad alteration in adaptive immune responsiveness.

Humoral responses were assessed using a binding antibody multiplex assay (BAMA) to quantify IgG subclass responses against HIV antigens. Levels of IgG1 and IgG3 targeting p24, p17, and gp120 remained stable across all timepoints (SF16), indicating that BCG vaccination did not measurably affect HIV-specific antibody responses in this cohort.

### Routine clinical follow-up reveals no major differences between study participants and non-participants

To assess whether the observed reduction in baseline immune activity was accompanied by differences in routine clinical outcomes, we used longitudinal follow-up data from the Swiss HIV Cohort Study (SHCS). BELIEVE participants were originally recruited from the SHCS-derived Systems-X cohort. This allowed comparison of the 55 BELIEVE study participants (Believer) with 751 non-participants (Non-believer) over a median follow-up of 41.2 (CI: 40.7-41.6) months after BCG vaccination or the matched reference date [30] (Table 1).

**Table 1.** Participant characteristics and HIV RNA burden by group.

|  | Believer | Non-believer |
| --- | --- | --- |
|  | (n = 55) | (n = 751) |
| **Peak HIV RNA, log10 copies/mL** |  |  |
| Median [IQR] | 5.13 [4.74, 5.54] | 5.16 [4.70, 5.60] |
| **Sex** |  |  |
| Female | 6 (10.9%) | 188 (25.0%) |
| Male | 49 (89.1%) | 563 (75.0%) |
| **Age at day 0, years** |  |  |
| Median [IQR] | 55.0 [49.0, 60.0] | 57.0 [50.0, 63.0] |
| **Mode of transmission** |  |  |
| Heterosexual | 13 (23.6%) | 284 (37.8%) |
| IDU | 2 (3.6%) | 37 (4.9%) |
| MSM | 39 (70.9%) | 379 (50.5%) |
| Unknown | 1 (1.8%) | 21 (2.8%) |
| Blood products | 0 (0.0%) | 9 (1.2%) |
| Hemophilia | 0 (0.0%) | 1 (0.1%) |
| IDU/Sexual | 0 (0.0%) | 13 (1.7%) |
| Other | 0 (0.0%) | 7 (0.9%) |
| **CD4 nadir, cells/μL** |  |  |
| Median [IQR] | 193 [141, 252] | 184 [82, 259] |
| **ART episodes, n** |  |  |
| Median [IQR] | 5.00 [3.50, 6.00] | 6.00 [4.00, 7.00] |
| **Cumulative ART duration, years** |  |  |
| Median [IQR] | 19.17 [18.32, 20.39] | 20.00 [18.35, 22.34] |
| **HIV RNA AUC = 0 after day 0** |  |  |
| Yes | 32 (58.2%) | 470 (62.6%) |
| No | 23 (41.8%) | 275 (36.6%) |
| **Time-normalized HIV RNA AUC after day 0, among participants with AUC > 0** |  |  |
| Median [IQR] | 0.18 [0.11, 0.28] | 0.27 [0.18, 0.52] |

Abbreviations: ART, antiretroviral therapy; AUC, area under the curve; IDU, injecting drug use; IQR, interquartile range; MSM, men who have sex with men

Longitudinal HIV RNA measurements remained largely suppressed in both groups, with similar group-level trajectories over follow-up (Fig. 5a). Consistently, time-normalized HIV RNA area under the curve (AUC) values were concentrated at or near 0 and did not differ between Believer and Non-believer participants (Fig. 5b). We next assessed clinical events grouped according to predefined SHCS clinical categories, including outcomes such as cardiovascular, metabolic, liver, kidney, and bone-related events. Overall, 146 clinical events were recorded in Non-believer, affecting 109 individuals (14.5%), compared with 12 events in Believer, affecting 9 individuals (16.4%). Clinical event rates normalized to person-years of follow-up were comparable across clinical categories (SF17a). Disease-related events, including infection-related and neoplastic conditions, showed a similar pattern. In total, 50 disease events were recorded in Non-believer, affecting 39 individuals (5.2%), compared with 4 events in Believer, affecting 4 individuals (7.3%). When grouped into neoplasia, infection-related events, and other diseases, event rates varied between categories but no obvious group-specific pattern was observed (SF17b).

**Figure 5.**
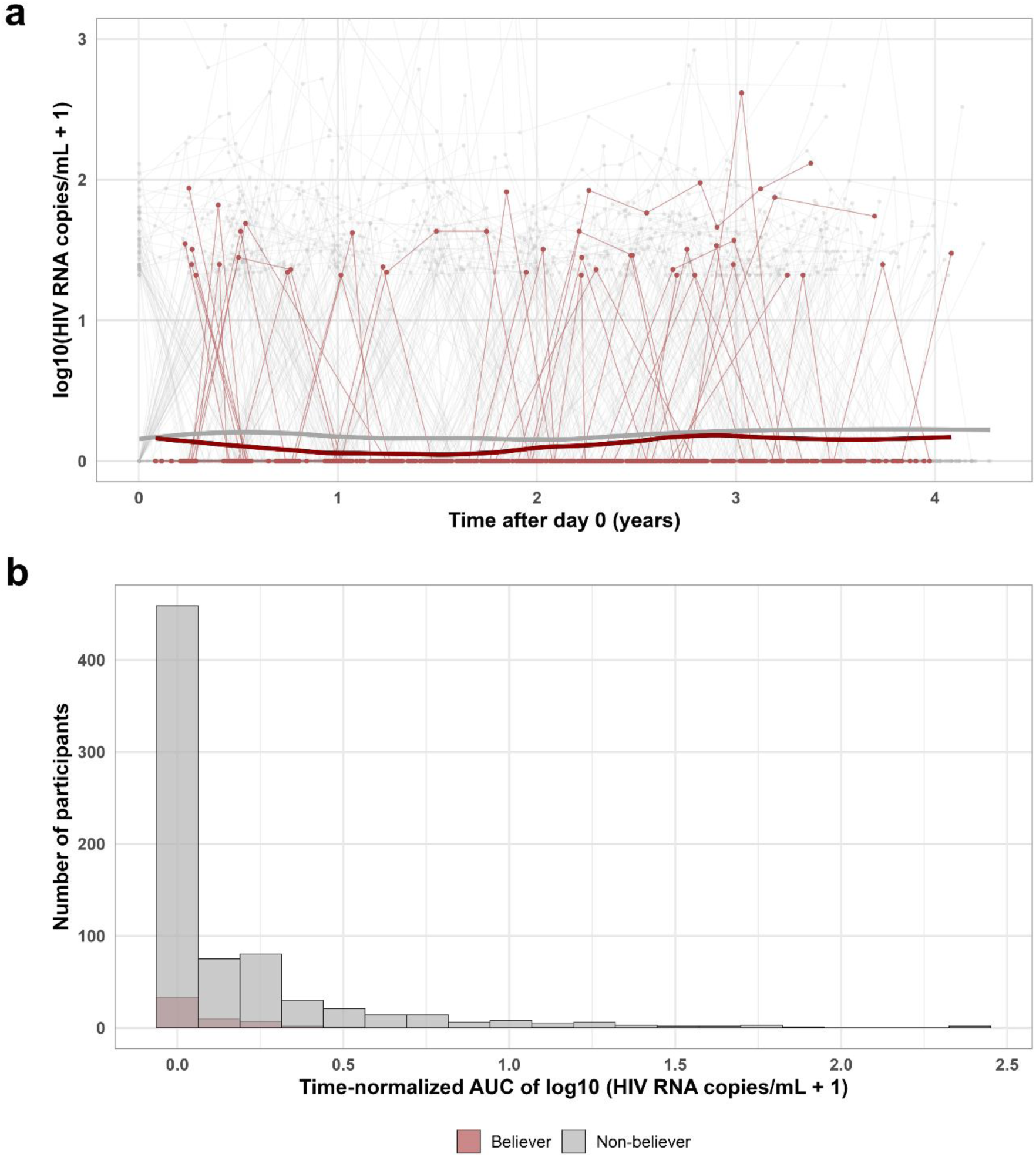
Longitudinal HIV RNA measurements in study participants and non-participants. a) Longitudinal HIV RNA measurements after BCG vaccination or the corresponding reference date in study participants (Believer) and non-participants (Non-believer). Values are shown as log10(HIV RNA copies/mL + 1). Thin lines and points represent individual participants. Thick lines represent group-specific locally estimated scatterplot smoothing (LOESS) curves fitted to the longitudinal measurements. b) Distribution of time-normalized HIV RNA area under the curve (AUC) after day 0 in Believer and Non-believer participants. AUCs were calculated by trapezoidal integration of log10(HIV RNA copies/mL + 1) over time and divided by the interval between the first and last available measurements. An AUC of 0 indicates that all HIV RNA measurements contributing to the AUC were recorded as 0. Statistics: Believer (n = 55) were compared with Non-believer (n = 751). Among participants with at least two HIV RNA measurements after day 0, HIV RNA burden was summarized per individual as the time-normalized trapezoidal AUC of log10(HIV RNA copies/mL + 1) and compared between groups using a Wilcoxon rank-sum test. Data were derived from routine clinical measurements.

Together, these data extend the clinical safety observations reported by West et al. [27] and suggest that the immunological shifts observed after BCG vaccination were not accompanied by obvious adverse patterns in descriptive routine clinical follow-up.

## Discussion

In this longitudinal study of BCG vaccination in ART-treated PWH, two findings stand out. First, BCG was associated with a coordinated reduction in tonic interferon-linked immune activity across functional, proteomic, transcriptional, and chromatin analyses. Second, selected immune functions remained preserved, including stimulus-induced cytokine production and Fc-dependent cytotoxicity. Together, these findings suggest that BCG reshaped innate immune activity toward lower baseline activation while maintaining responses to secondary stimulation and antibody-coated targets.

The immune configuration observed after BCG vaccination partially parallels immune states described in non-progressive lentiviral infection. Natural SIV hosts rapidly resolve chronic interferon-associated activation despite persistent viremia, and some elite-controller cohorts combine comparatively low systemic inflammation with strong HIV-specific cytotoxic responses, although the underlying mechanisms and interferon profiles remain heterogeneous [7–11,13]. However, interferon-associated and innate activation profiles remain heterogeneous among elite controllers, and some cohorts retain evidence of substantial innate activation despite spontaneous viral control [31]. BELIEVE extends these observations by showing in a controlled vaccination setting that lower interferon-linked activity can emerge together with preserved inducible and Fc-dependent function. This does not establish mechanistic equivalence to natural control or tolerance, but it supports the broader principle that reduced constitutive activation need not entail generalized loss of antiviral responsiveness. This interpretation is also consistent with the SHCS observations that motivated the trial, in which asymptomatic Mtb infection was associated with lower HIV viral set points, selective reduction of interferon-associated transcriptional programs, and altered HIV-specific antibody responses [14–16].

The reduction in baseline IFN-γ-associated activity is particularly noteworthy in the context of treated HIV infection. Persistent immune activation and interferon signaling can remain detectable despite long-term viral suppression on ART and have been linked to ongoing immune dysregulation and non-AIDS comorbidities [2,32,33]. Against this background, lower tonic interferon activity may represent attenuation of a chronically activated immune state. The reduction in basal IFN-γ production was accompanied by molecular changes, including decreased interferon-stimulated gene expression, lower IRF- and STAT-associated motif activity, and reduced IFN-γ-related pathway activity in plasma proteomics, most prominently at day 90. Together, these concordant observations support a coordinated shift in immune state rather than an isolated functional change.

Effector functions were not uniformly affected. PBMC-mediated direct cytotoxicity and unstimulated NK cell IFN-γ production decreased, whereas stimulated cytokine production remained inducible and ADCC was preserved. This pattern indicates differential regulation of constitutive, direct, and Fc-dependent effector pathways.

The absence of a classical circulating trained immunity phenotype provides additional context for these findings. Although BCG is often associated with enhanced heterologous myeloid responsiveness [20], we observed little evidence of sustained augmentation of monocyte cytokine production. Instead, reduced IFN-γ-linked activity was apparent across several immune compartments. IFN-γ can establish durable transcriptional and chromatin programs in macrophages [34], and recent work emphasizes that BCG-induced heterologous effects arise through interactions between adaptive and innate immunity [35,36]. The altered interferon environment may therefore have contributed to the distinct myeloid phenotype observed here. Circulating myeloid cells increased, but this was accompanied by reduced HLA-DR expression and unchanged heterologous TNF-α responses rather than evidence of enhanced activation. Although this relationship cannot be established directly from our data, the findings reinforce the concept that BCG-induced effects depend on host immune context and may differ between healthy individuals and PWH [25,37–40].

The preservation of Fc-dependent effector function may have particular relevance. Broadly neutralizing antibodies can engage Fcγ receptor-expressing effector cells and promote the elimination of HIV-infected targets in addition to neutralizing free virus, although the contribution of Fc-mediated mechanisms varies across antibodies and experimental systems [41–44]. Although the rituximab-Raji assay does not model HIV-specific antibody recognition, it shows that Fc-dependent cellular killing remained functional despite reduced direct cytotoxicity. In vitro studies have likewise shown that BCG exposure can modulate Fc-dependent NK cell activity [45,46]. These findings provide a rationale for directly testing whether BCG-induced immune changes influence Fc-dependent antibody activity in HIV-specific systems.

The immunological changes occurred without broad alterations in the adaptive parameters examined and without serious BCG-related safety events in the parent BELIEVE trial, despite frequent local reactions [27]. Longer-term follow-up likewise did not identify major differences in virological or clinical outcomes, although these analyses were limited by low event numbers and their observational nature. These observations are consistent with selective recalibration of circulating immune activity without evidence of broad functional impairment.

This study has several limitations. Sampling at days 90 and 180 may have missed early transient responses, including short-lived myeloid activation. Because the immune analyses relied primarily on within-participant longitudinal comparisons without a parallel unvaccinated immunological control group, time-dependent changes unrelated to vaccination cannot be fully excluded. The exploratory single-cell multiomic analysis used pooled samples from only three donors per timepoint and did not permit donor-level statistical inference. Cytotoxicity and ADCC were assessed in whole-PBMC co-cultures, preserving intercellular interactions but limiting attribution to altered per-cell NK cell function versus effector-cell composition. CD56 depletion nevertheless supported a substantial NK cell contribution. Finally, the cohort comprised clinically stable, virologically suppressed PWH in a low-TB-endemic setting, which may limit generalizability to other immune states and populations. Future studies should assess tissue-resident immunity, HIV-specific Fc-mediated responses, and the clinical relevance of changes in tonic interferon activity.

Overall, BCG vaccination in ART-treated PWH was associated with a coordinated reduction in tonic interferon-linked and direct cytotoxic activity while preserving inducible cytokine responses and antibody-dependent killing. These findings identify a selective reorganization of immune function shaped by the host immune context and provide a rationale for testing its relevance to Fc-dependent antibody interventions in HIV-specific systems.

## Methods

### Clinical specimens

Cryopreserved peripheral blood mononuclear cells (PBMCs) and plasma samples from 55 people with HIV (PWH) participating in the Swiss HIV Cohort Study (SHCS) [47] and the clinical trial reported by West et al. [27] were analyzed. The study was approved by the Ethikkommission Zürich (BASEC ID 2021-01481). All participants provided written informed consent prior to enrollment. The trial is registered at ClinicalTrials.gov (NCT05004038).

Timepoints were defined relative to the time of BCG vaccination, with day 0 representing baseline, and follow-up samples were obtained from all participants 90 and 180 days after vaccination. Participants received intradermal BCG (AJ Vaccines; *Mycobacterium bovis* Danish strain 1331; 2-8 × 10⁵ CFU).

### PBMC stimulation and flow cytometry staining

Cryopreserved PBMCs were thawed in a 37 °C water bath and washed in pre-warmed R10 medium (RPMI 1640 supplemented with 10% FBS, 1% L-glutamine, and 1% penicillin-streptomycin if required). Cells were stimulated for 4 h at 37 °C and 5% CO_2_ in the presence of Brefeldin A with one of the following stimuli (200 µL per well): PMA/Ionomycin (1:1000 Cell Activation Cocktail), heat-killed *Mycobacterium tuberculosis* (approx. MOI 5, 5×10^6^ bacteria per well), LPS (from *E. coli* K12; 50 ng/mL), β-glucan (from *S. cerevisiae*; 5 µg/mL), or R10 medium as unstimulated control.

For extracellular staining, cells were incubated for 15 min at room temperature (RT) with TruStain FcX (1:20) and Zombie NIR (1:500), washed with FACS buffer (PBS + 2% FBS, 2 mM EDTA, 0.1% sodium azide), and stained with antibody mixes (Supplementary Table 1) for 25 min at RT in the dark.

After fixation and permeabilization with the Cyto-Fast™ Fix/Perm Buffer Set, intracellular antibodies (Supplementary Table 1) were added and incubated for 25 min at RT.

Finally, cells were fixed in 2% paraformaldehyde and stored at 4 °C until acquisition on a Cytek Aurora 5L spectral flow cytometer (Cytometry Facility, University of Zurich).

### High-dimensional flow cytometry analysis

Flow cytometry standard (FCS) files were processed using *Cytolution* (Cytolytics, Germany; pipeline version 1.2.0) for automated data cleaning, normalization, and batch correction. Normalized and arcsinh-transformed FCS files were exported for downstream analysis. Processed FCS files were analyzed in R using diffcyt [48]. Cells were first over-clustered with FlowSOM. Clusters were then aggregated into biologically coherent metaclusters by meta-clustering and manual curation. Manual gating within *Cytolution* was used to quantify specific cell subpopulations or extract marker-level parameters (e.g. proportions of cytokine-positive cells).

### Compartment-level immune correlation analysis

To assess changes in immune co-variation over time, compartment-level meta-correlation networks were constructed from flow cytometry-derived readouts at day 0, day 90, and day 180. Included features comprised major immune-cell proportions, lymphoid and myeloid cytokine-positive cell frequencies, unstimulated NK phenotype frequencies, and myeloid HLA-DR median expression. At each timepoint, pairwise Spearman correlations were calculated between all included features using complete observations. Correlations were retained as feature-level edges if |r| > 0.4 and the Benjamini-Hochberg FDR-adjusted p value was <0.05. Features were assigned to myeloid cells, T cells, B cells, or NK(T) cells, and feature-level edges were collapsed into compartment-level networks. Inter-compartment connectivity was quantified as edge density, defined as the number of significant correlations between two compartments divided by the total number of possible feature pairs between them. Edge widths in the meta-networks represent edge density and were plotted on a shared scale across timepoints.

### Cytotoxicity assay

K562, ACH-2, and A3.01 cells were cultured in R10 medium. 24 h before the assay, ACH-2 and A3.01 cells were stimulated for 22-24 h with 0.5 ng/mL TNF-α [49]. Target cells were labeled with CellTrace™ CFSE (2.5 µM) for 20 min at 37 °C, followed by quenching in medium for 5 min at RT, and seeded (10’000 cells per well) in 96-well plates overnight.

Cryopreserved PBMCs were thawed, washed, and counted before being added to target cells (190’000 PBMCs per well; E:T ratio 19:1) for 4 h at 37 °C and 5% CO_2_. Cells were then stained in PBS containing 3 µM DRAQ7™ for 10 min at 37 °C. Samples were directly analyzed on a CytoFLEX S flow cytometer (Beckman Coulter). Cytotoxicity was calculated by subtracting spontaneous target cell death in target cell-only wells from the proportion of dead target cells in co-culture conditions.

### Antibody-dependent cellular cytotoxicity (ADCC) assay

Cryopreserved PBMCs were thawed, washed, and rested overnight in R10 medium. Raji cells were cultured in R10 medium, labeled with CellTrace™ CFSE (1 µM) for 10 min at 37 °C, followed by quenching in medium for 5 min at RT, and seeded (5’000 cells per well) in 96-well plates. PBMCs were added at 195’000 cells per well (E:T ratio 39:1). Cells were co-cultured for 6 h at 37 °C and 5% CO_2_ in the presence or absence of rituximab (0.5 µg/mL). Cells were then stained in PBS containing 3 µM DRAQ7 for 10 min at 37 °C and directly analyzed on a CytoFLEX S flow cytometer (Beckman Coulter). Specific rituximab-dependent killing was calculated by subtracting the proportion of dead target cells in rituximab-negative conditions from the corresponding rituximab-positive co-culture conditions.

### Single-cell multiome (ATAC + Gene Expression) profiling

Single-cell multiome profiling was performed at the Functional Genomics Center Zurich (FGCZ). PBMCs from three donors at each timepoint were pooled separately for day 0 and day 90 and co-cultured with K562 cells as described in the cytotoxicity assay. CD56⁺ cells were enriched using CD56 MicroBeads (Miltenyi Biotec), and nuclei were extracted using the 10x Genomics Nuclei Isolation Kit following the manufacturer’s protocol. Approximately 16’000 nuclei per sample were loaded onto the 10x Genomics Chromium Controller, and single-cell multiome libraries were prepared using the Chromium Next GEM Single Cell Multiome ATAC + Gene Expression kit (v3.1, 10x Genomics). Libraries were sequenced on an Illumina NovaSeq X Plus.

Raw sequencing data were processed with Cell Ranger ARC (v2.0.2, 10x Genomics) using the GRCh38.p13 human reference genome (GENCODE Release 42). For the gene expression modality, ambient RNA contamination was removed with CellBender, and low-quality cells, erythrocyte contamination, and non-NK populations were filtered. Cell type classification was performed using scGate [50] with curated PBMC models, followed by semi-supervised batch correction and integration with STACAS [51]. NK cells were subsetted for downstream analysis, and clusters were identified using Seurat. Label transfer from a published NK cell reference atlas [29] was performed to assess correspondence with established NK cell subtypes.

Gene expression patterns were assessed at the single-cell level using selected marker genes and curated gene modules. Positive cluster markers were identified using Seurat FindAllMarkers. Average expression and module scores for curated NK cell programs and selected MSigDB Hallmark gene sets were calculated using Seurat functions and summarized by cluster and condition, with differences between day 90 and day 0 evaluated descriptively.

For the ATAC modality, peak count matrices and fragment files were processed with Signac. Chromatin accessibility data were normalized using TF-IDF (term frequency-inverse document frequency) and reduced via LSI (latent semantic indexing). Quality control metrics included TSS enrichment, nucleosome signal, and fragment length distribution. Transcription factor motifs from JASPAR2022 were added using the hg38 reference genome, and motif activity was quantified using chromVAR. Within RNA-defined clusters 0 and 2, motif activity differences between day 90 and day 0 were ranked using logistic regression in Seurat, adjusting for ATAC fragment counts and TSS enrichment. This cell-level analysis was used for exploratory motif ranking only. Median chromVAR deviation scores were summarized by cluster and condition, and chromatin accessibility at selected loci was visualized using Signac coverage plots. Multimodal integration was performed using Weighted Nearest Neighbors (WNN) [52] to jointly analyze transcriptional and chromatin accessibility states. Because samples from the three donors were pooled within each timepoint, comparisons between day 0 and day 90 were interpreted descriptively and no donor-level inferential statistical testing was performed.

### Proteomics data processing and analysis

Proteomic data were acquired as previously described by Ryan et al. [53] at the FGCZ, using the same sample preparation workflow and instrumentation. Protein data matrices (626 protein groups measured) were exported and used as input for downstream analyses in R. Sample quality was assessed by principal component analysis and by evaluating potential sample-handling artifacts using platelet contamination, erythrocyte contamination, and coagulation indices, as described by Geyer et al. [54]. Samples identified as quality control outliers were excluded before downstream analysis.

Proteins quantified in at least 70% of samples were retained (481 protein groups remained). Intensities were transformed as log2(x + 1), and missing values were imputed using a left-censored minimum-probability approach implemented in imputeLCMD (impute.MinProb, q = 0.01). Median normalization was applied across samples, followed by batch correction for measurement plate effects using ComBat from the sva package.

Differential protein abundance across timepoints was assessed using linear models with empirical Bayes moderation implemented in limma. Repeated measurements were accounted for using duplicateCorrelation, with participant as the blocking factor. Contrasts compared day 90 with day 0 and day 180 with day 0. Gene set enrichment analysis was performed using fgseaMultilevel from the fgsea package. Protein groups containing multiple UniProt identifiers were expanded and mapped to gene symbols. When more than one protein group mapped to the same gene, the entry with the largest absolute moderated t-statistic was retained. Gene symbols were then ranked by the moderated t-statistic from the corresponding limma contrast. For targeted analyses, a predefined subset of seven immune-related MSigDB Hallmark pathways [55] (v2025.1.Hs) was analyzed, with FDR correction applied across the included pathways separately for each contrast. Normalized enrichment scores and leading-edge genes were used for interpretation and visualization.

### In vitro HIV infection assay and TZM-bl reporter readout

Cryopreserved PBMCs were thawed, washed, and stimulated for 48 h at 37 °C and 5% CO_2_ in R10 medium supplemented with 10 U/mL IL-2, and 5 µg/mL phytohemagglutinin (PHA). Cells were seeded at 4×10⁶ cells/mL in a 24-well plate during stimulation.

Following stimulation, cells were washed and resuspended in culture R10 medium supplemented with 50 U/mL IL-2 at 2×10⁶ cells/mL and infected with HIV-1. Virus stocks of YU2 or NL4-3 (TCID_50_-titrated) were added to achieve multiplicities of infection (MOI) of 0.1 or 0.01 (YU2) and 0.01 or 0.001 (NL4-3) in a final infection volume of 100 µL per well. Cells were incubated with virus for 24 h at 37 °C and 5% CO_2_, washed with PBS, and cultured for an additional 5 days in 200 µL culture medium.

Supernatants were harvested and used to infect TZM-bl reporter cells. TZM-bl cells were seeded at 1×10⁴ cells per well in white 96-well plates in 100 µL DMEM containing 10% FBS and 1% L-glutamine. PBMC supernatant (50 µL) was added and cells were incubated for 48 h.

Infection was quantified using a luciferase reporter assay (Promega). Cells were lysed with 50 µL 1X Passive Lysis Buffer and luciferase substrate was added according to the manufacturer’s instructions. Luminescence was measured using a GloMax Navigator luminometer and reported as relative light units (RLU).

### T cell proliferation assay

Cryopreserved PBMCs were thawed and counted, and 1×10⁶ cells per condition were labeled with the proliferation dye Tag-it Violet (2.5 µM) for 20 min in a 37 °C water bath, followed by quenching in R10 medium for 5 min at RT. Labeled cells were resuspended in R10 medium, seeded in 24-well plates (2 mL/well), and cultured for 4 days at 37 °C and 5% CO_2_ under unstimulated conditions or in the presence of T-Activator CD3/CD28 beads (0.5 beads per cell).

After incubation, cells were harvested, washed with PBS, and subjected to extracellular staining in FACS buffer for 25-30 min at room temperature (RT) using antibodies against CD3, CD4, CD8, and a viability dye. Cells were then fixed and permeabilized using the eBioscience FoxP3 Transcription Factor Staining Buffer Set (Thermo Fisher Scientific) according to the manufacturer’s instructions, followed by intracellular staining for Ki-67 and TCF1 in permeabilization buffer. After washing, cells were resuspended in FACS buffer and analyzed on a CytoFLEX S flow cytometer (Beckman Coulter). T cell proliferation was quantified as the proportion of cells showing dilution of Tag-it Violet.

### HIV-1 binding antibody multiplex assay (BAMA)

Biotinylated target proteins were coupled to different regions of MagPlex-Avidin Microspheres as previously described [56]. In brief, 320 nM protein in PBS (JR-FL gp120, p24, p17 or human serum albumin as control) was coupled to 1.25×10^6^ MagPlex-Avidin Microspheres for 1 h at RT. Beads were washed three times in PBS containing 1% BSA (PBS-BSA 1%) and stored at −20 °C until use.

To measure the HIV-specific antibody response of the participants, plasma was heat-inactivated for 1 h at 56 °C and diluted 1:100 in PBS-BSA 1%. Beads coupled to different antigens were mixed and adjusted to 20 beads of each antigen-coupled bead region per µL. 50 µL of diluted plasma was incubated with 50 µL of the bead mix for 1 h at RT. After washing three times with PBS-BSA 1%, beads were incubated for 1 h with detector antibodies for IgG1 (Mouse anti-human IgG1-PE) or IgG3 (Mouse anti-human IgG3-PE) at a final concentration of 1:500 in PBS-BSA 1%. Beads were washed three times with PBS-BSA 1% and median fluorescence intensity (MFI) was measured on the FlexMap 3D reader (Luminex Corporation). A minimum of 50 bead reads per antigen was acquired.

### Normalization of fluorescence intensity signal obtained in BAMA

Raw MFI values were normalized in R (version 4.4.1; brms package), separately for IgG1 and IgG3, to correct for plate-to-plate variability, subtract assay background, and account for the limits of quantification of the assay.

Positive binding controls (pools of plasma from people with chronic HIV infection) were included on every plate, and a Bayesian hierarchical regression was used to rescale each plate’s measurements onto a common reference scale, thereby removing inter-plate variability. Per-antigen assay background was estimated from HIV-negative healthy-donor plasma included as controls and subtracted from each sample, and a per-antigen lower limit of detection (LOD) was derived from this background. Normalized values were then restricted to the quantifiable range of the assay, between the LOD and the instrument’s upper limit (10^5^ MFI), and used for all subsequent antibody-response analyses.

### Statistical analysis

Differential abundance of cell type proportions and differential state analysis of marker expression in flow cytometry data were performed using diffcyt-DA-GLMM and diffcyt-DS-LMM, respectively [48], with timepoint as a fixed effect and participant as a random intercept. Planned contrasts compared day 90 and day 180 with baseline (day 0), and p values were adjusted for multiple testing using the FDR. Longitudinal analyses of other immune readouts were carried out using linear mixed-effects models implemented in lme4, with timepoint as a fixed effect and participant as a random intercept. For stimulation assays, stimulant, timepoint, and their interaction were included as fixed effects, excluding the unstimulated condition. Readouts were analyzed as log2 fold changes relative to baseline or, for selected stimulation assays, relative to the corresponding unstimulated condition. Estimated marginal means and planned contrasts were obtained using emmeans, with p values adjusted for multiple testing using the FDR. For supplementary analyses of raw cytotoxicity and ADCC values, day 90 and day 180 were compared with day 0 using paired t tests, with FDR adjustment across the two longitudinal contrasts.

Routine clinical follow-up analyses were performed separately from the longitudinal immune analyses. For individuals with at least two measurements after BCG vaccination or the matched reference date, HIV RNA burden was summarized as the time-normalized trapezoidal AUC of log10(HIV RNA copies/mL + 1) and compared between BELIEVE participants and non-participants using a Wilcoxon rank-sum test. Clinical and disease events were summarized as rates per 100 person-years of follow-up. Numbers of events per participant were compared between groups within each event category using Wilcoxon rank-sum tests, with FDR adjustment across categories.

All statistical analyses were performed in R (versions 4.4.1-4.5.2).

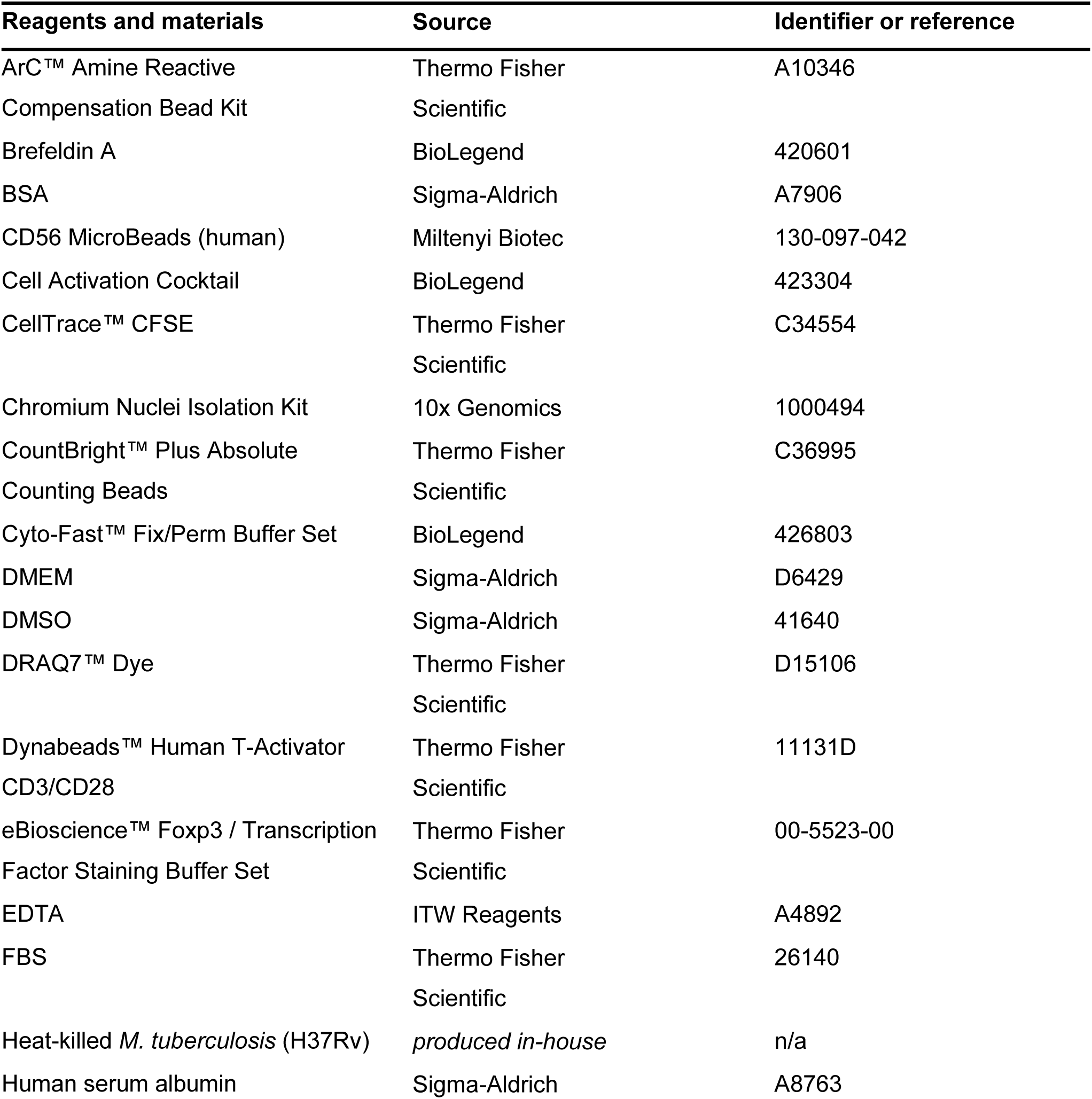
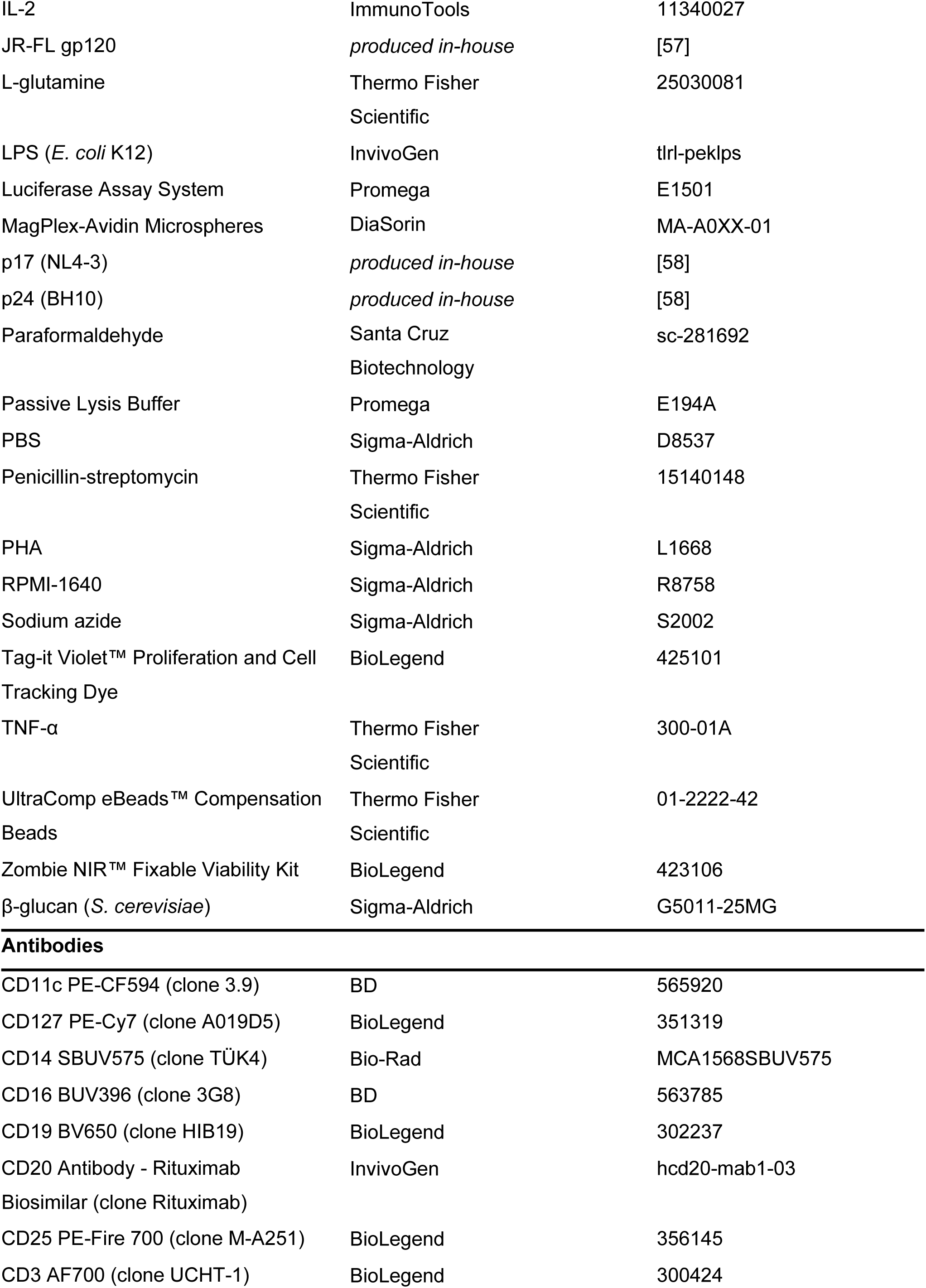
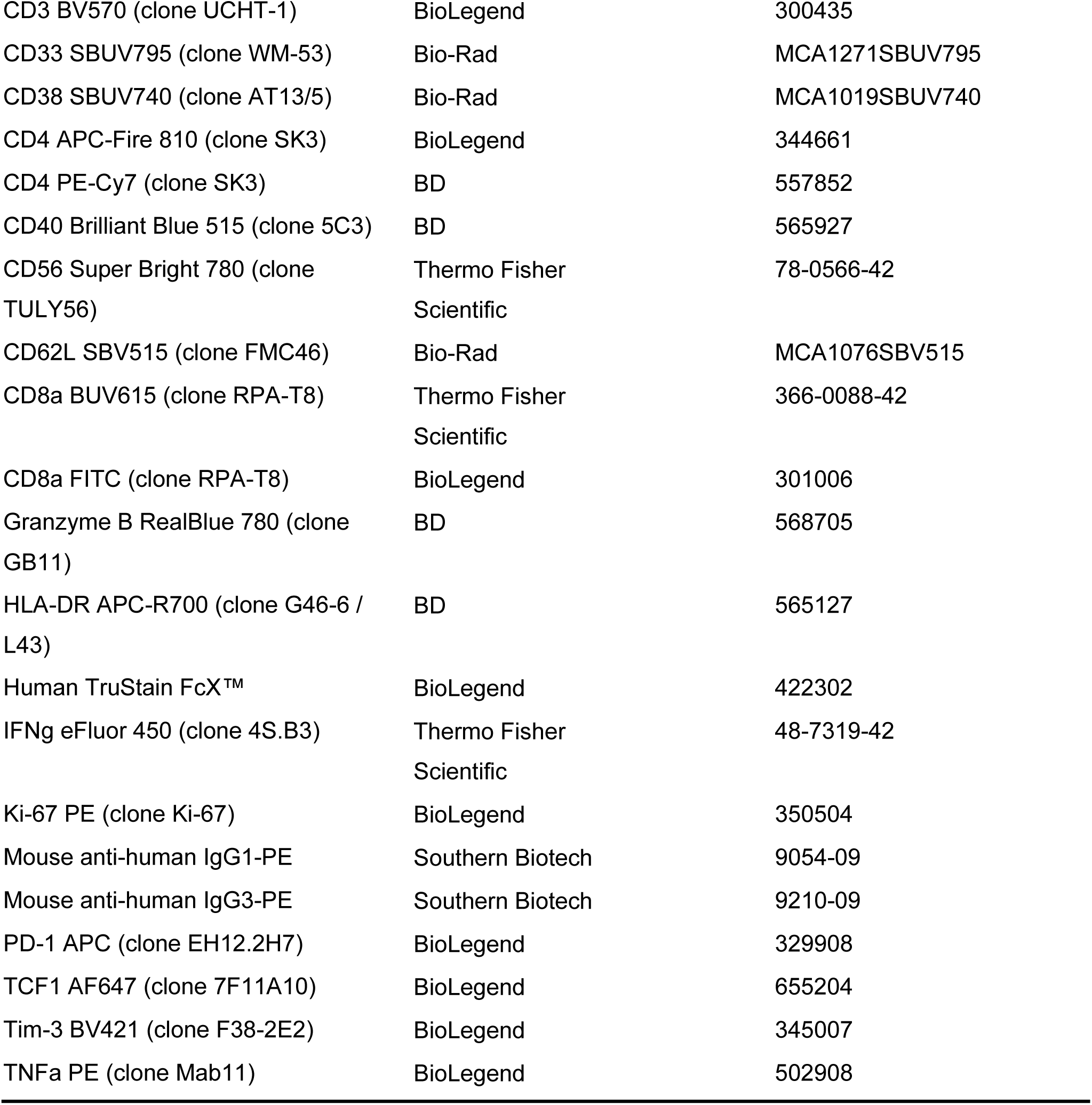

## Acknowledgments

We would like to thank the study participants and the Swiss HIV Cohort Study for their contributions and support. We would also like to thank all the study nurses, data managers, and physicians who were involved in data collection. Flow cytometry was performed in part using equipment of the Cytometry Facility, University of Zurich. Flow cytometry data were analyzed using *Cytolution* (Cytolytics GmbH, Tübingen, Germany) as part of the data processing workflow. Single-cell multiome profiling and proteomics analyses were performed at the Functional Genomics Center Zurich (FGCZ), University of Zurich and ETH Zurich. We thank Tatiane Gorski (Cytometry Facility), Can Pinar (Cytolytics GmbH), Paolo Nanni (FGCZ, proteomics), Nikolai Püllen (FGCZ, single-cell sequencing), and Paul Gueguen (FGCZ, bioinformatics) for their assistance with data analysis and technical support. The authors used large language models (ChatGPT and Claude) to assist with language editing, improve clarity, and support the preparation and refinement of analysis code. The tools were not used to generate scientific conclusions or interpret results. All code, analyses, and manuscript content were reviewed, verified, and edited by the authors, who take full responsibility for the final manuscript. Parts of this work were presented at the Conference of the European AIDS Clinical Society (October 2025, Paris; ePoster) and the Congress of the European Society of Clinical Microbiology and Infectious Diseases (April 2026, Munich; oral presentation).

## Author contributions

Conceptualization: JN, CD, EW, DLB; Methodology: JN, CD, EW, DLB; Investigation: CD, TKT, DR, MS; Formal analysis: CD, TKT, LB, BD, MZ; Validation: BD, MZ, KK, JN; Data curation: CD, TKT, MZ, KK, BE, MS; Project administration: CD, JN; Visualization: CD, BD, LB; Resources: JN, HFG, AT; Supervision: JN, HFG, AT; Funding acquisition: JN, HFG; Writing - original draft: CD, JN; Writing - review & editing: All authors.

## Data availability

The Swiss HIV Cohort Study (SHCS) established in 1988 collects clinical data and biological samples from people living with HIV in Switzerland. All participants have provided written informed consent; however, strict confidentiality rules apply. Specifically, the SHCS is obligated to ensure that individuals cannot be re-identified from shared data, which prevents the public release of detailed participant-level data (https://www.shcs.ch/for-researchers/open-data-statement-shcs/). For projects that require participant-level data variables, a proposal needs to be submitted to the Scientific Board of the SHCS (https://www.shcs.ch/for-researchers/who-can-submit/).

## Supplementary materials

**Supplementary Figure 1.**
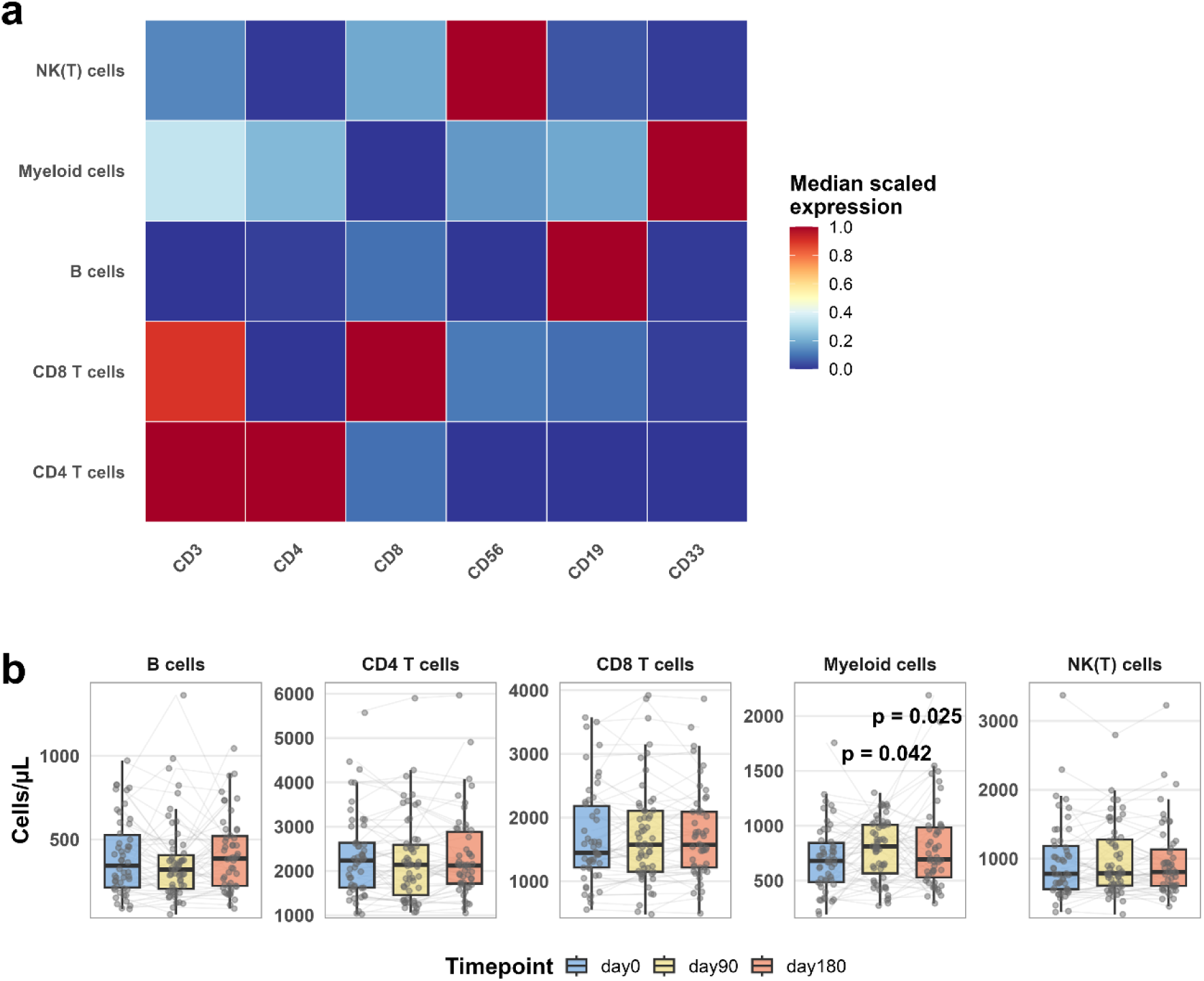
Definition of immune cell populations by lineage marker expression and absolute cell counts. a) Heatmap showing median scaled expression of lineage markers (x-axis) used to define major immune cell populations (y-axis) in PBMCs. b) Boxplots showing absolute cell numbers across timepoints, with lines and points representing individual participants (n = 55). Absolute counts were estimated by scaling flow cytometry-derived cell proportions to corresponding clinical leukocyte counts. Statistics: Longitudinal changes were assessed using linear mixed-effects models, with planned contrasts of day 90 and day 180 vs. day 0. P values were FDR-adjusted; only adjusted p < 0.05 are indicated. Data were derived from flow cytometry.

**Supplementary Figure 2.**
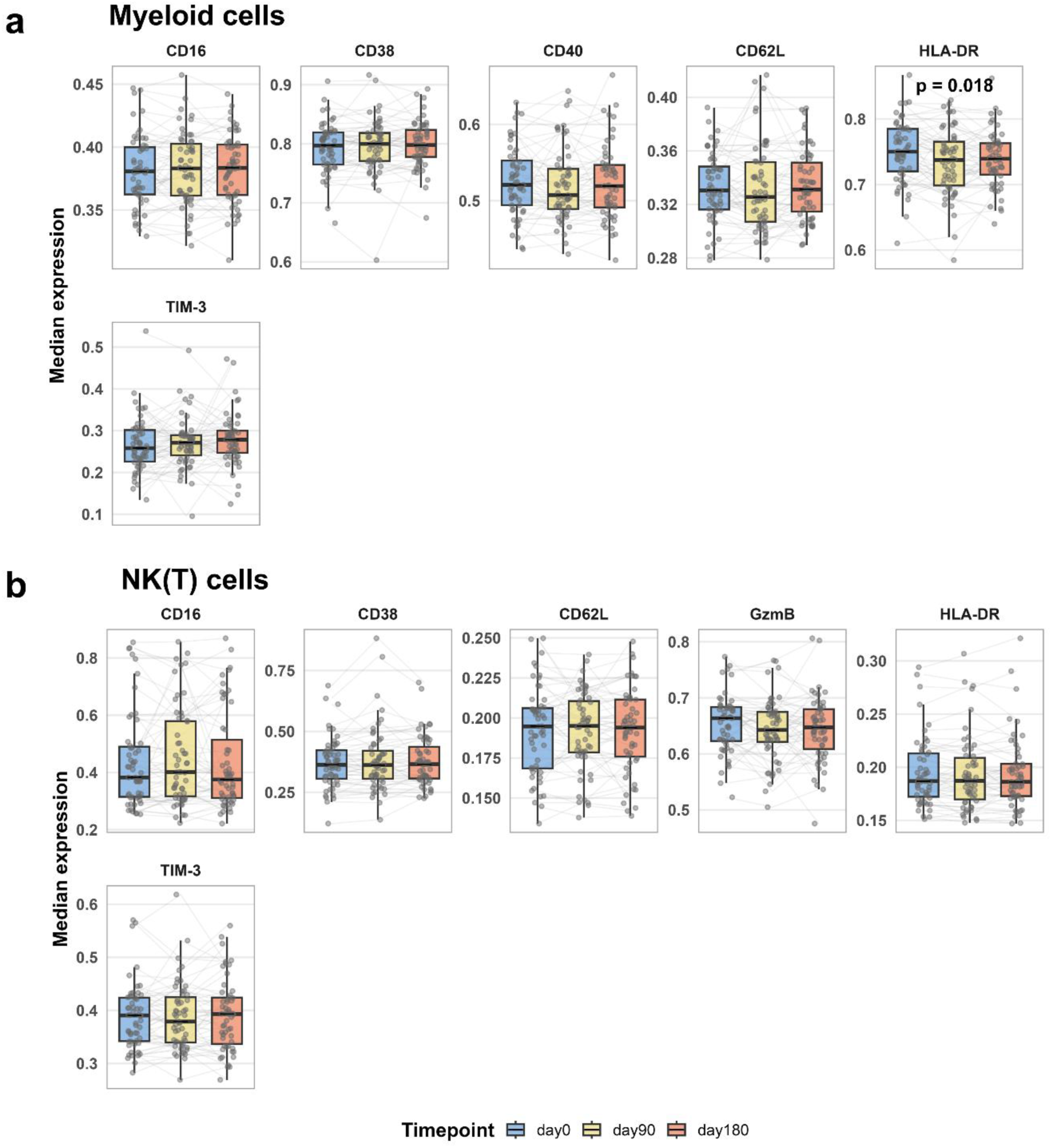
Expression of selected markers in myeloid and NK(T) cells. a) Boxplots showing median expression of selected markers in CD33^+^ myeloid cells across timepoints. b) Boxplots showing median expression of selected markers in CD56^+^ cells across timepoints. Statistics: Differential state was assessed using diffcyt-DS-LMM, with planned contrasts of day 90 and day 180 vs. day 0. P values were FDR-adjusted; only adjusted p < 0.05 are indicated. Data were derived from flow cytometry.

**Supplementary Figure 3.**
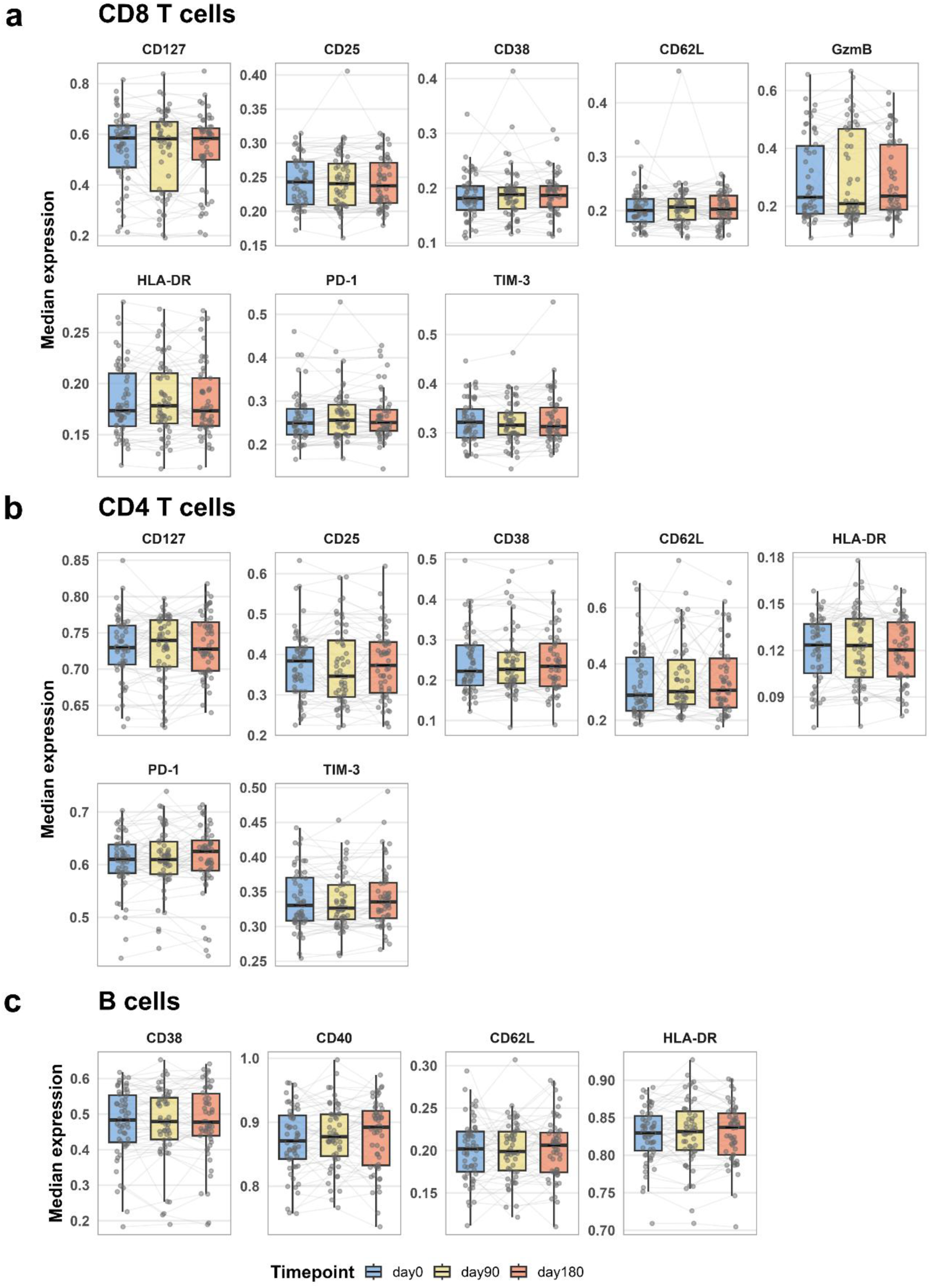
Expression of selected markers in B and T cells. a) Boxplots showing median expression of selected markers in CD8^+^ T cells across timepoints. b) Boxplots showing median expression of selected markers in CD4^+^ T cells across timepoints. c) Boxplots showing median expression of selected markers in B cells across timepoints. Statistics: Differential state was assessed using diffcyt-DS-LMM, with planned contrasts of day 90 and day 180 vs. day 0. P values were FDR-adjusted; only adjusted p < 0.05 are indicated. Data were derived from flow cytometry.

**Supplementary Figure 4.**
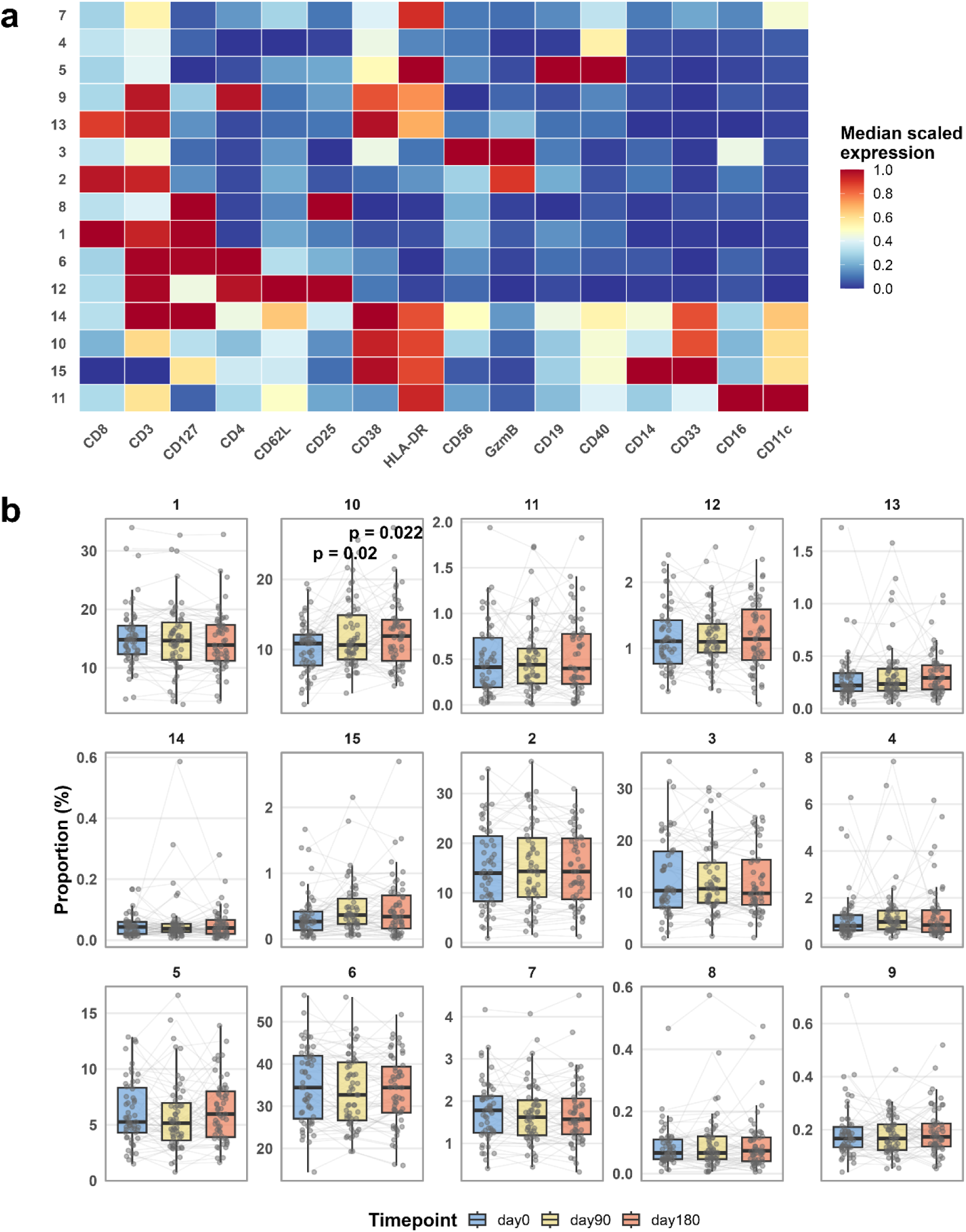
Automated clustering of PBMCs based on high-dimensional marker expression. a) Heatmap showing median scaled expression of markers included in clustering (x-axis), used to automatically define 15 clusters (y-axis) in PBMCs. b) Boxplot of cluster proportions across timepoints, with lines and points representing individual participants. Statistics: Differential abundance was assessed using diffcyt-DA-GLMM, with planned contrasts of day 90 and day 180 vs. day 0. P values were FDR-adjusted; only adjusted p < 0.05 are indicated. Data were derived from flow cytometry.

**Supplementary Figure 5.**
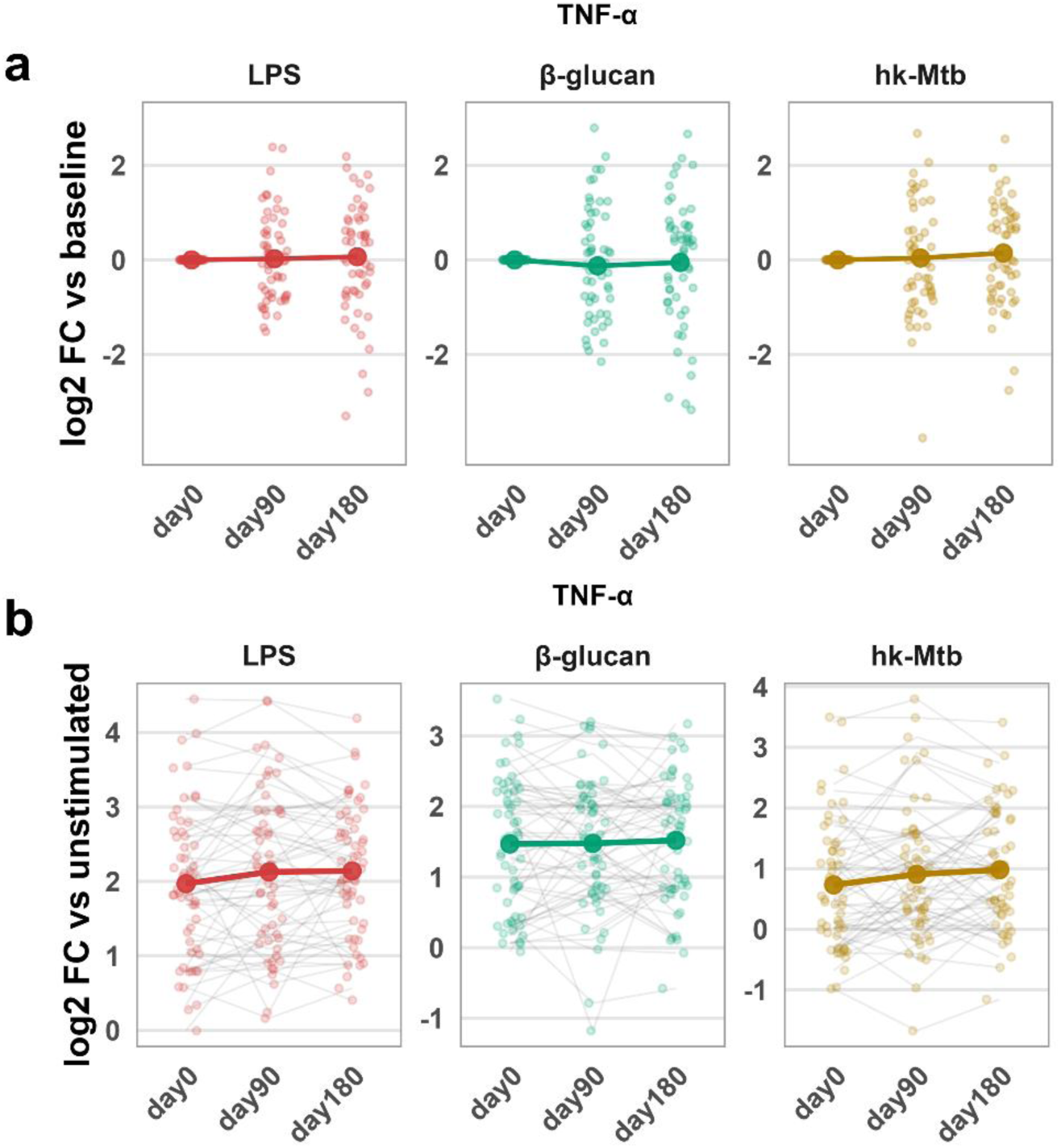
No evidence for trained immunity-like responses in myeloid cells after BCG vaccination. a) Log2 fold-changes relative to baseline (day 0) of TNF-α-producing CD33^+^ myeloid cells following stimulation with LPS, β-glucan, or heat-killed *M. tuberculosis* (hk-Mtb). Lines represent estimated marginal means and points represent individual participants (n = 55). b) Log2 fold-changes relative to the corresponding unstimulated condition of TNF-α-producing CD33^+^ myeloid cells following stimulation with LPS, β-glucan, or hk-Mtb. Thick line represents estimated marginal means and points with connecting thin lines represent individual participants (n = 55). Statistics: Longitudinal changes in stimulation assays were assessed using linear mixed-effects models, with planned contrasts of day 90 and day 180 vs. day 0. P values were FDR-adjusted; only adjusted p < 0.05 are indicated. Data were derived from flow cytometry.

**Supplementary Figure 6.**
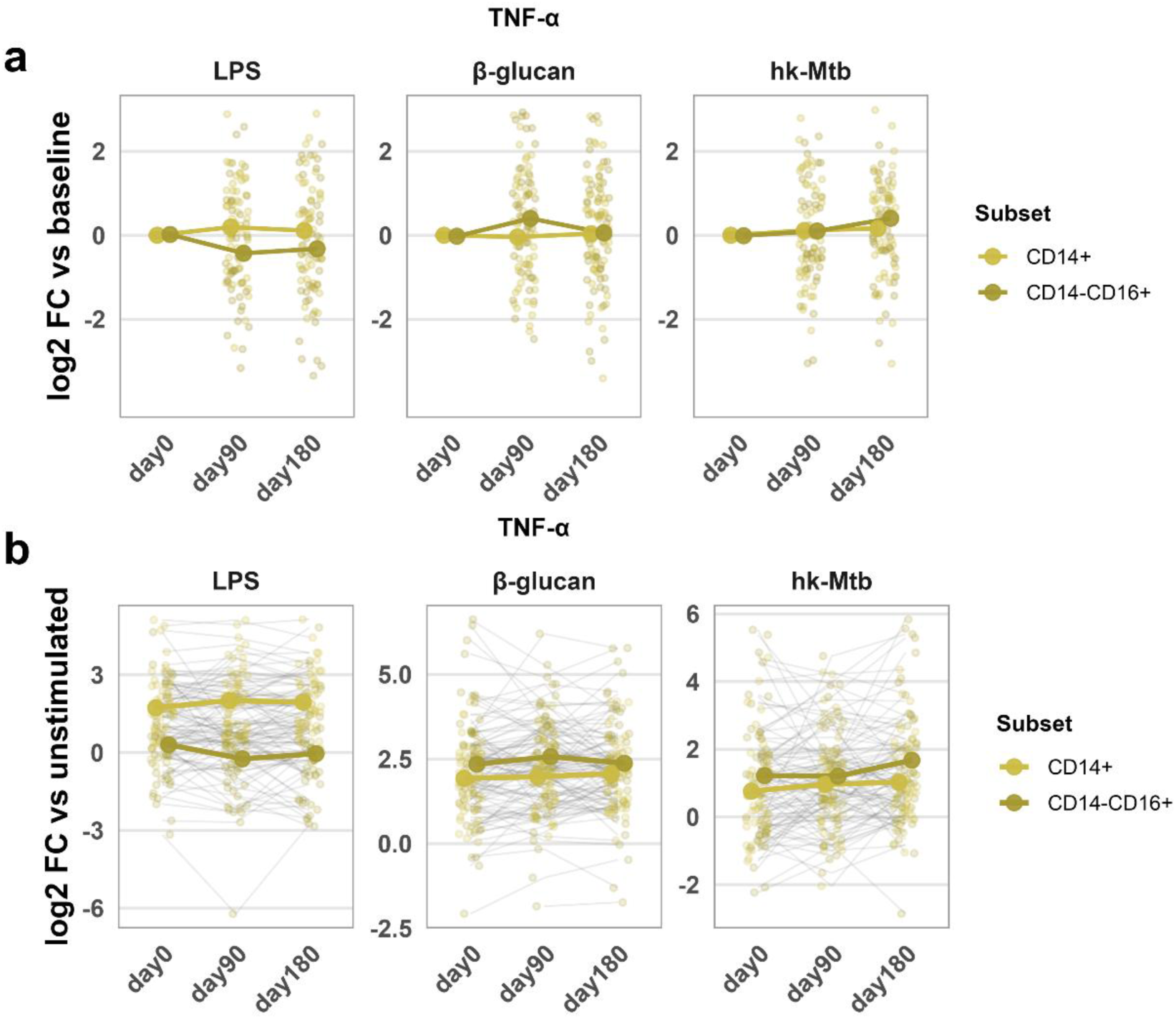
Monocyte subsets do not exhibit enhanced TNF-α responses after BCG vaccination. a) Log2 fold-changes relative to baseline (day 0) of TNF-α-producing CD14^+^ and CD14^−^CD16^+^ monocyte subsets following stimulation with LPS, β-glucan, or heat-killed *M. tuberculosis* (hk-Mtb). Lines represent estimated marginal means and points represent individual participants (n = 55). b) Log2 fold-changes relative to the corresponding unstimulated condition of TNF-α-producing CD14^+^ and CD14^−^ CD16^+^ monocyte subsets following stimulation with LPS, β-glucan, or hk-Mtb. Thick line represents estimated marginal means and points with connecting thin lines represent individual participants (n = 55). Statistics: Longitudinal changes in stimulation assays were assessed using linear mixed-effects models, with planned contrasts of day 90 and day 180 vs. day 0. P values were FDR-adjusted; only adjusted p < 0.05 are indicated. Data were derived from flow cytometry.

**Supplementary Figure 7.**
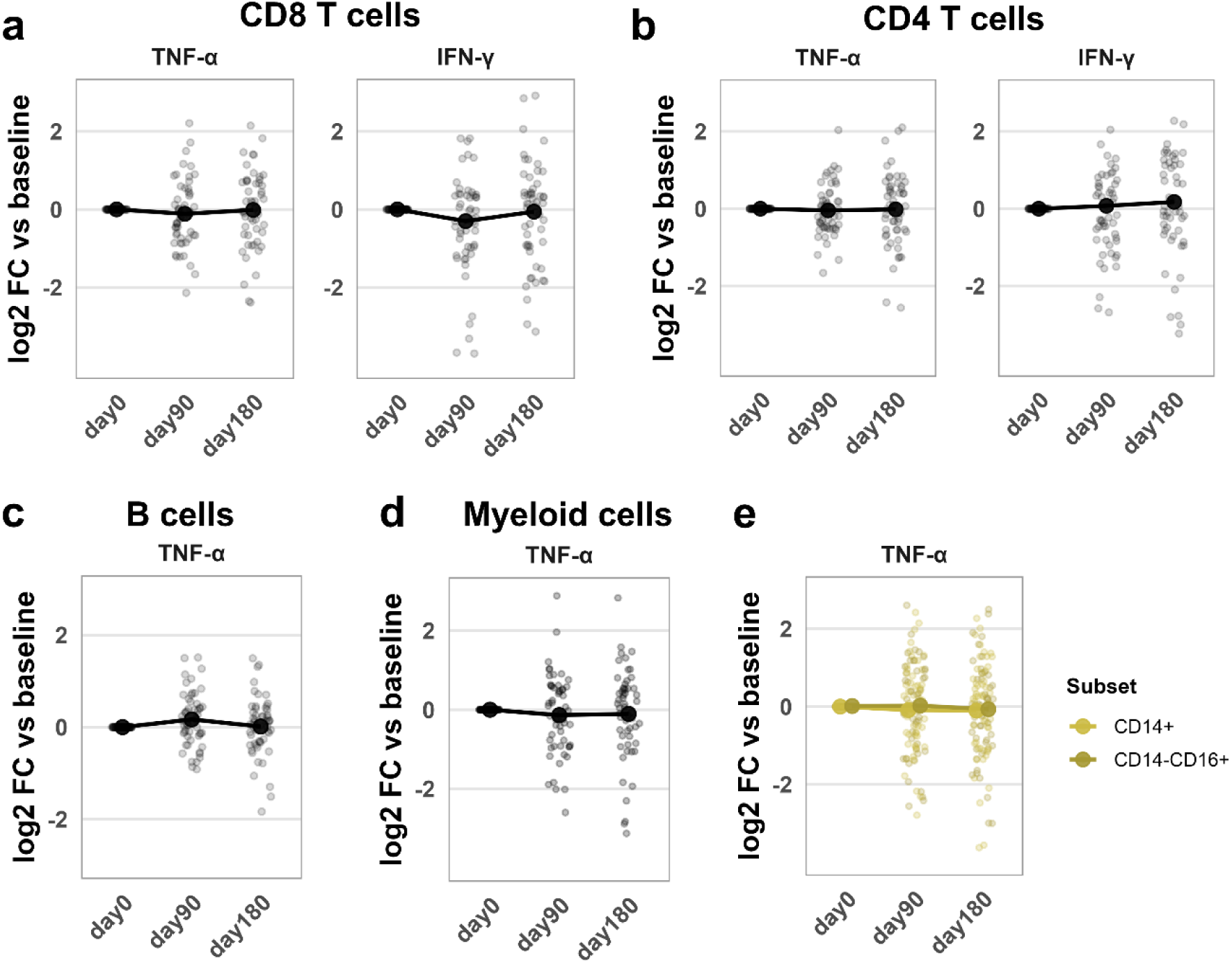
Limited longitudinal changes in cytokine production across other major immune-cell populations. a) Log2 fold-changes relative to baseline (day 0) of TNF-α- and IFN-γ-producing CD8 T cells following 4 h brefeldin A treatment under unstimulated conditions. Lines represent estimated marginal means and points represent individual participants (n = 55). b) Log2 fold-changes relative to baseline (day 0) of TNF-α- and IFN-γ-producing CD4 T cells following 4 h brefeldin A treatment under unstimulated conditions. Lines represent estimated marginal means and points represent individual participants (n = 55). c) Log2 fold-changes relative to baseline (day 0) of TNF-α-producing B cells following 4 h brefeldin A treatment under unstimulated conditions. Lines represent estimated marginal means and points represent individual participants (n = 55). d) Log2 fold-changes relative to baseline (day 0) of TNF-α-producing CD33^+^ myeloid cells following 4 h brefeldin A treatment under unstimulated conditions. Lines represent estimated marginal means and points represent individual participants (n = 55). e) Log2 fold-changes relative to baseline (day 0) of TNF-α-producing CD14^+^ and CD14^−^CD16^+^ monocyte subsets following 4 h brefeldin A treatment under unstimulated conditions. Lines represent estimated marginal means and points represent individual participants (n = 55). Statistics: Longitudinal changes were assessed using linear mixed-effects models, with planned contrasts of day 90 and day 180 vs. day 0. P values were FDR-adjusted; only adjusted p < 0.05 are indicated. Data were derived from flow cytometry.

**Supplementary Figure 8.**
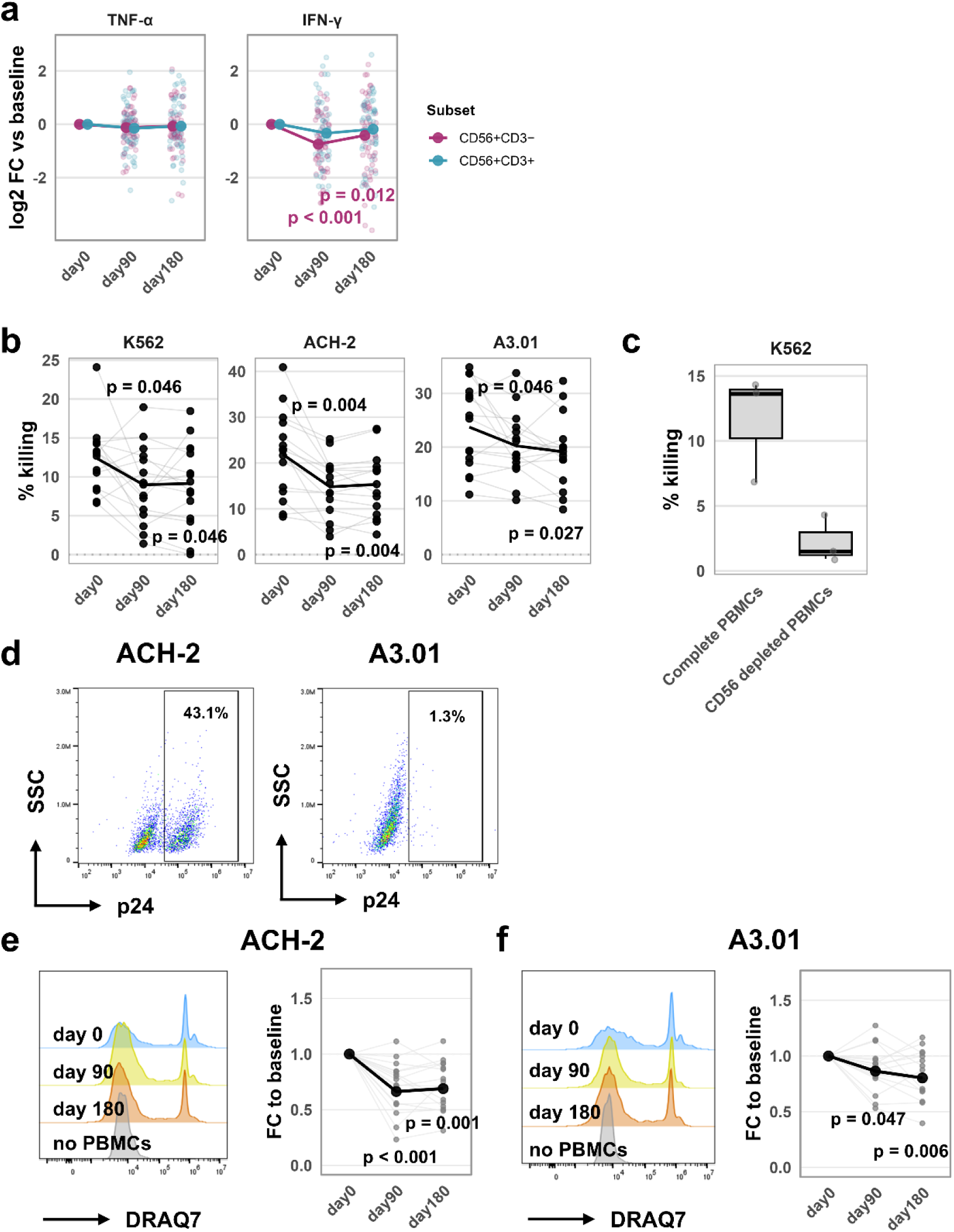
NK cell cytokine production and target-cell killing. a) Log2 fold-changes relative to baseline (day 0) of TNF-α- and IFN-γ-producing NK (CD3^−^) and NKT (CD3^+^) cells following 4 h brefeldin A treatment under unstimulated conditions. Lines represent estimated marginal means and points represent individual participants (n = 55). b) Percentage of killed target cells after 4 h co-culture of PBMCs with K562, ACH-2, or A3.01 target cells across timepoints. Each point represents an individual participant (n = 15). Specific killing was calculated by subtracting spontaneous target-cell death in target-cell-only wells from target-cell death after co-culture. c) Boxplots showing the percentage of killed K562 target cells after 4 h co-culture with either whole PBMCs or CD56-depleted PBMCs. Data shown are from three timepoint-matched participants. d) Representative intracellular staining showing HIV-1 p24 expression in ACH-2 and A3.01 cells. e) Representative flow cytometry histogram showing DRAQ7 staining used to quantify target-cell death after 4 h co-culture of ACH-2 cells with PBMCs. Fold-changes relative to day 0 are shown. Thick line represents estimated marginal means and points with connecting thin lines represent individual participants (n = 15). f) Representative flow cytometry histogram showing DRAQ7 staining used to quantify target-cell death after 4 h co-culture of A3.01 cells with PBMCs. Fold-changes relative to day 0 are shown. Thick line represents estimated marginal means and points with connecting thin lines represent individual participants (n = 15). Statistics: Raw specific-killing values in panel b were compared using paired t tests. Longitudinal changes in panels a, e, and f were assessed using linear mixed-effects models, with planned contrasts of day 90 and day 180 vs. day 0. P values were FDR-adjusted; only adjusted p < 0.05 are indicated. Data were derived from flow cytometry.

**Supplementary Figure 9.**
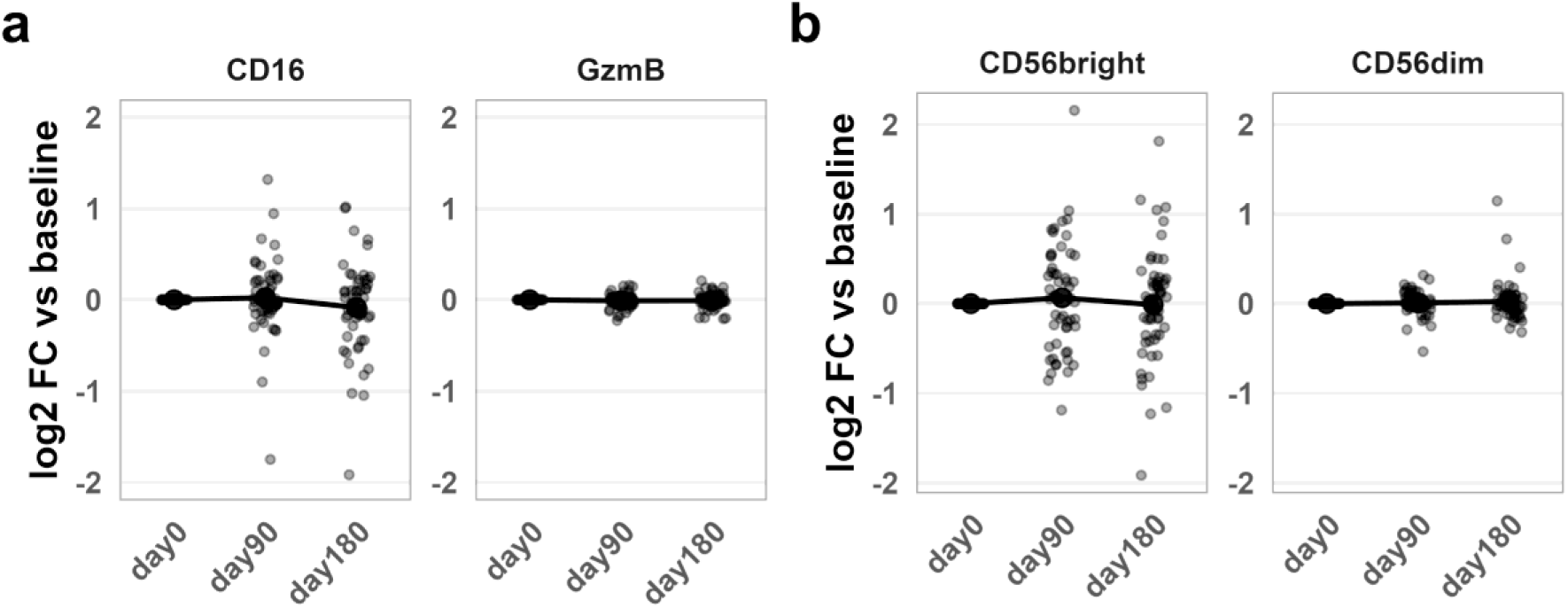
NK cell marker expression and phenotype distribution across timepoints. a) Log2 fold-changes relative to baseline (day 0) of CD16^+^ and GzmB^+^ CD56^+^ cells following 4 h brefeldin A treatment under unstimulated conditions. Lines represent estimated marginal means and points represent individual participants (n = 55). b) Log2 fold-changes relative to baseline (day 0) of CD56bright and CD56dim NK (CD3^−^) cells following 4 h brefeldin A treatment under unstimulated conditions. Lines represent estimated marginal means and points represent individual participants (n = 55). Statistics: Longitudinal changes were assessed using linear mixed-effects models, with planned contrasts of day 90 and day 180 vs. day 0. P values were FDR-adjusted; only adjusted p < 0.05 are indicated. Data were derived from flow cytometry.

**Supplementary Figure 10.**
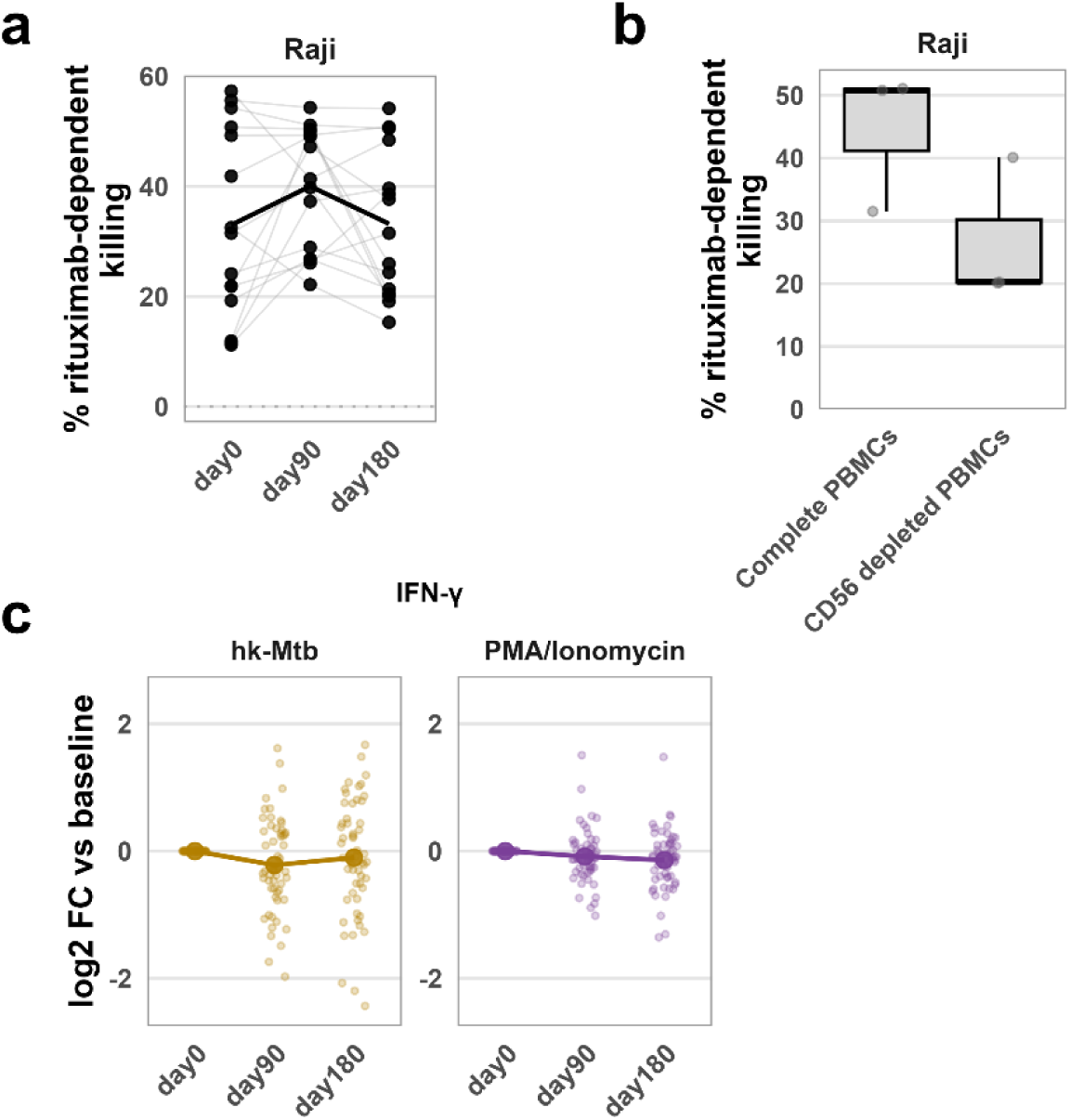
Antibody-dependent target-cell killing and stimulated NK cell cytokine responses. a) Percentage of rituximab-dependent killed target cells after 6 h co-culture of PBMCs with Raji target cells across timepoints. Each point represents an individual participant (n = 15). Specific killing was calculated by subtracting target-cell death in rituximab-negative conditions from that in the corresponding rituximab-positive conditions. b) Boxplots showing the percentage of killed Raji target cells after 6 h co-culture with either whole PBMCs or CD56-depleted PBMCs. Data shown are from three timepoint-matched participants. c) Log2 fold-changes relative to baseline (day 0) of TNF-α- and IFN-γ-producing CD56^+^ cells following 4 h brefeldin A treatment under heat-killed *M. tuberculosis* (hk-Mtb) and PMA/Ionomycin stimulation. Lines represent estimated marginal means and points represent individual participants. Statistics: Raw specific rituximab-dependent killing values in panel a were compared using paired t tests. Longitudinal changes in panel c were assessed using linear mixed-effects models, with planned contrasts of day 90 and day 180 vs. day 0. P values were FDR-adjusted; only adjusted p < 0.05 are indicated. Data were derived from flow cytometry.

**Supplementary Figure 11.**
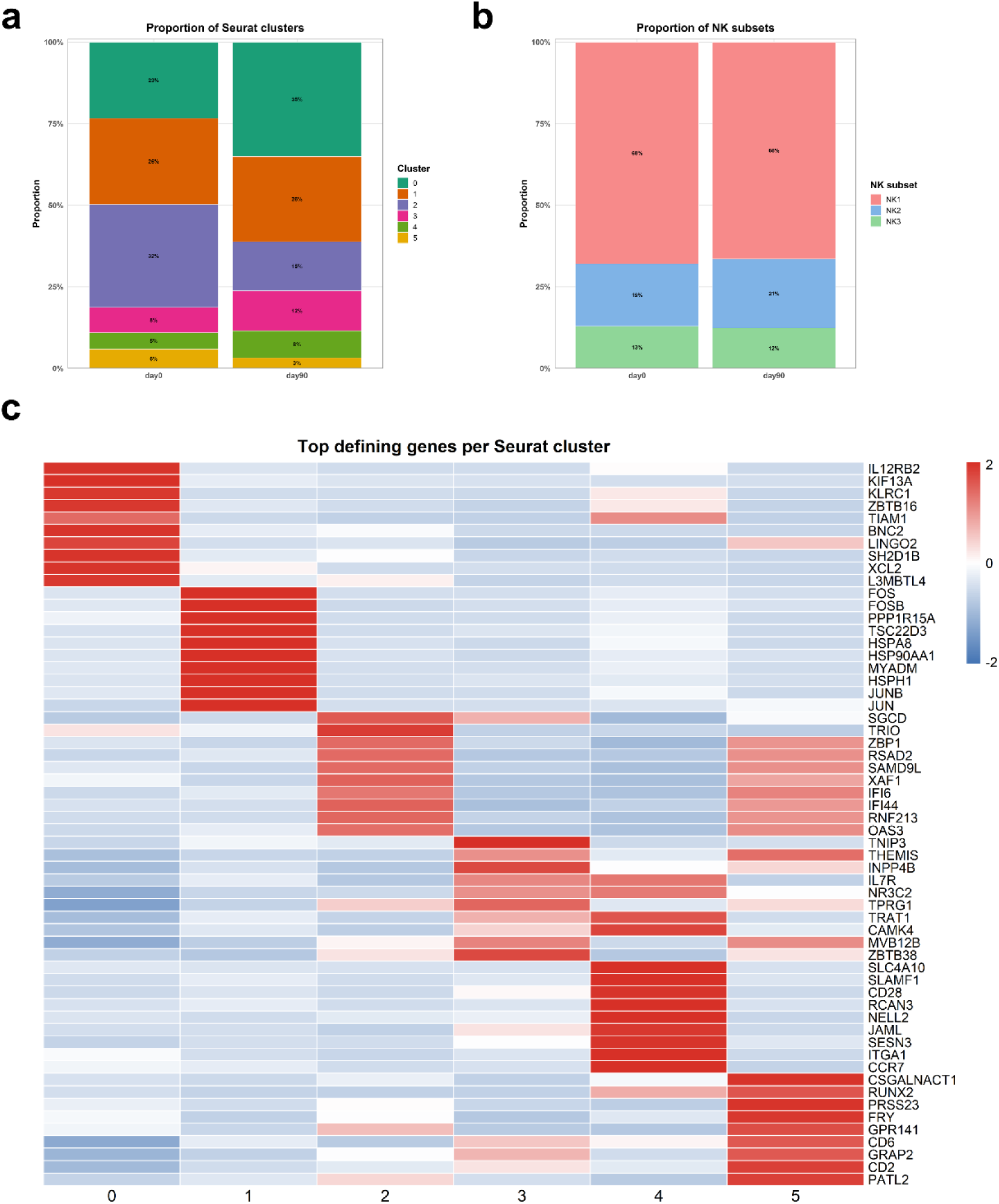
Single-cell RNA profiling of NK cells. a) Stacked bar plots showing proportions of RNA-based Seurat clusters (0-5) at day 0 and day 90. b) Stacked bar plots showing proportions of NK cell subsets (NK1, NK2, NK3) assigned by label transfer from a published NK cell reference atlas at day 0 and day 90. c) Heatmap showing row-scaled average expression of the top marker genes across Seurat clusters. Genes were selected based on differential expression and expression values were averaged per cluster. Statistics: Samples from three donors were pooled within each condition and timepoint; therefore, no donor-level inferential testing was performed, and results are shown descriptively. Single-cell RNA measurements were derived from single-cell multiome profiling.

**Supplementary Figure 12.**
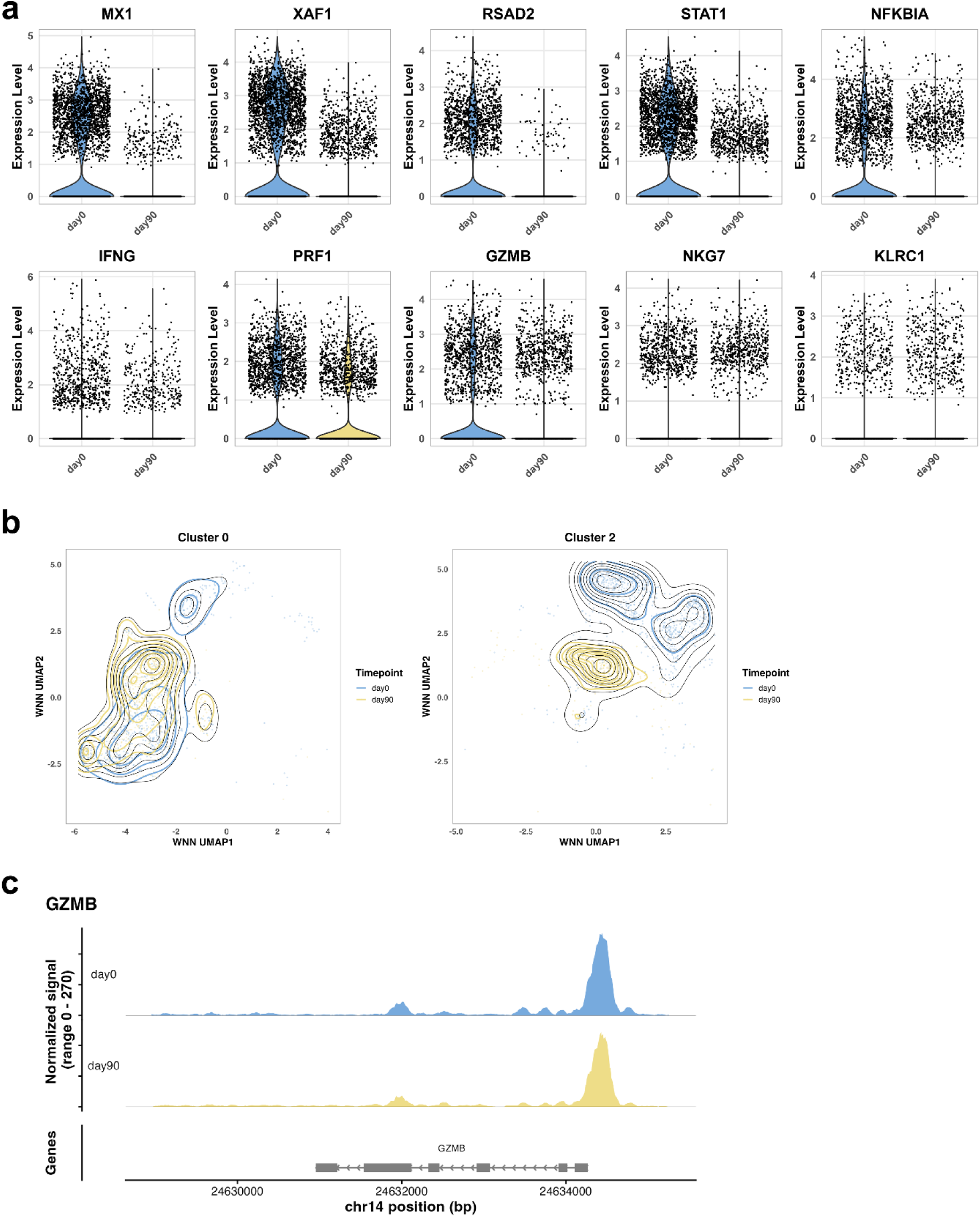
Transcriptional and chromatin features of NK-cell states across timepoints. a) Violin plots showing single-cell expression of selected interferon-stimulated and NK-associated genes at day 0 and day 90. Cells are grouped by timepoint, with each point representing an individual cell. b) Weighted nearest-neighbor (WNN) UMAP embedding of clusters 0 and 2 derived from integrated RNA and ATAC data. Cells are colored by timepoint (day 0 and day 90), with overlaid density contours indicating cell distribution within each condition. c) Chromatin accessibility coverage plot for the GZMB locus, showing aggregated ATAC signal at day 0 and day 90. Statistics: Samples from three donors were pooled within each condition and timepoint; therefore, no donor-level inferential testing was performed, and results are shown descriptively. Single-cell RNA and ATAC measurements were derived from single-cell multiome profiling.

**Supplementary Figure 13.**
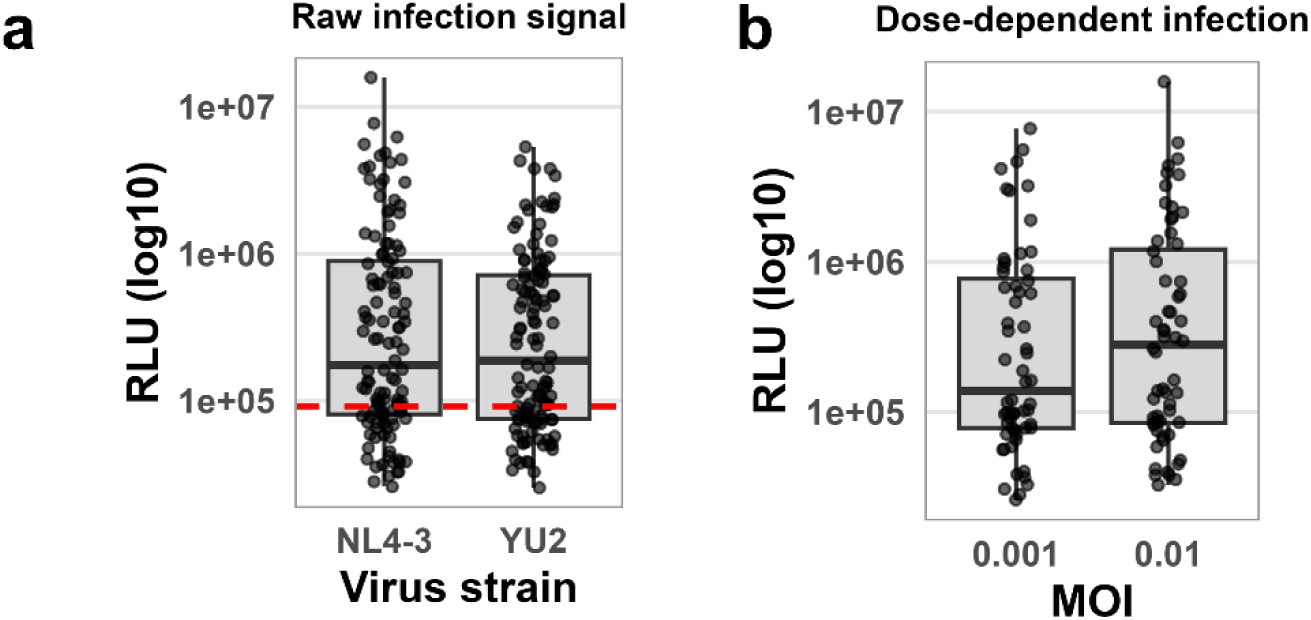
Validation of the in vitro HIV infection assay. a) Boxplots showing raw infection signals for HIV-1 strains NL4-3 and YU2 measured by TZM-bl luciferase activity in relative light units (RLU). Overlaid points represent individual participants (n = 20; three timepoints and two MOIs per strain). The red dashed line indicates the median background signal across two uninfected controls. b) Boxplots showing dose-dependent infection signal for HIV-1 NL4-3 at MOIs 0.001 and 0.01, with overlaid points representing individual participants. Statistics: No statistical testing was performed; data are shown descriptively. Data were derived from a TZM-bl reporter assay.

**Supplementary Figure 14.**
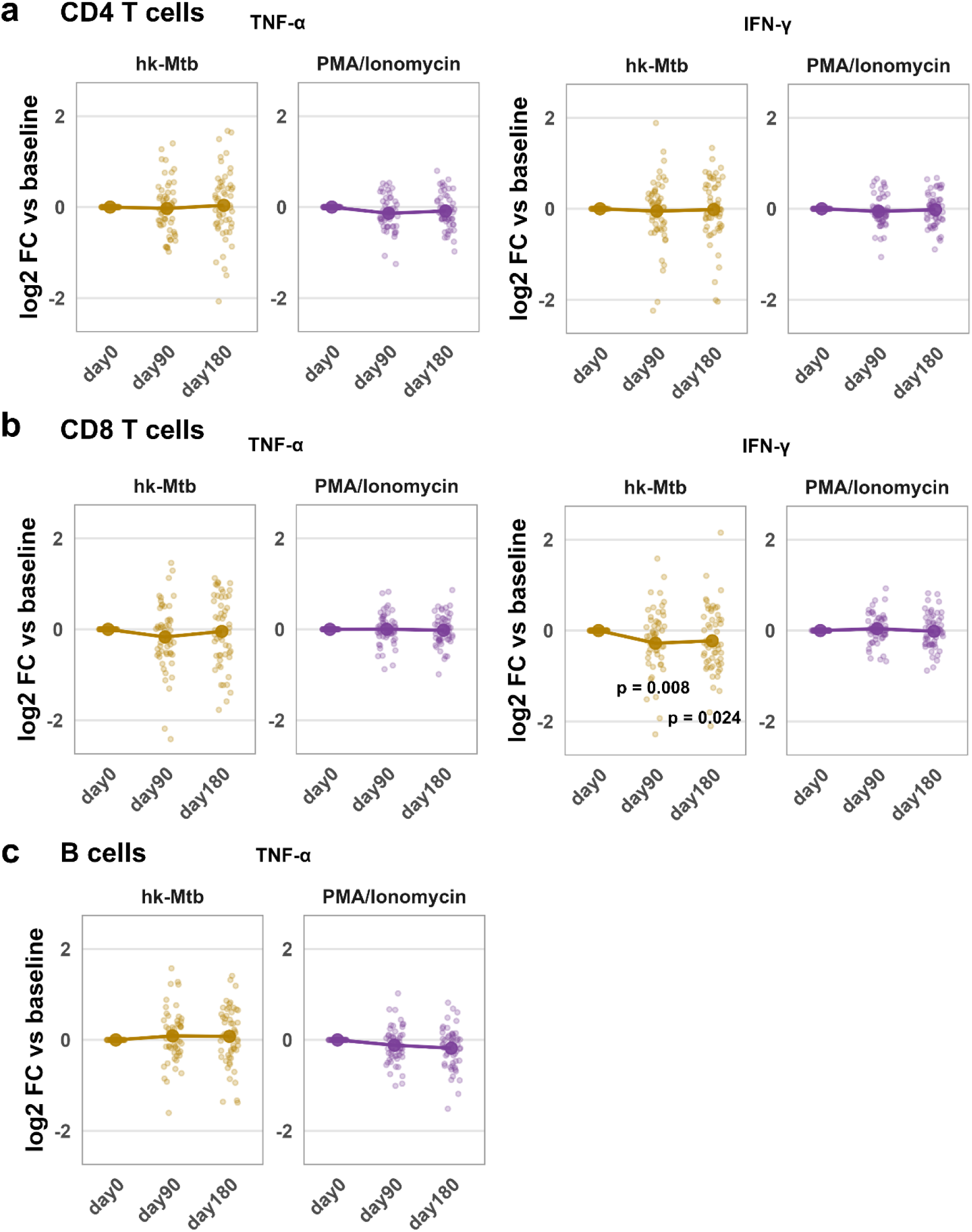
Cytokine production by lymphocyte subsets following stimulation. a) Log2 fold-changes relative to baseline (day 0) of TNF-α- and IFN-γ-producing CD4^+^ T cells following 4 h brefeldin A treatment under heat-killed *M. tuberculosis* (hk-Mtb) and PMA/Ionomycin stimulation. Lines represent estimated marginal means and points represent individual participants (n = 55). b) Log2 fold-changes relative to baseline (day 0) of TNF-α- and IFN-γ-producing CD8^+^ T cells following 4 h brefeldin A treatment under hk-Mtb and PMA/Ionomycin stimulation. Lines represent estimated marginal means and points represent individual participants (n = 55). c) Log2 fold-changes relative to baseline (day 0) of TNF-α-producing B cells following 4 h brefeldin A treatment under hk-Mtb and PMA/Ionomycin stimulation. Lines represent estimated marginal means and points represent individual participants (n = 55). Statistics: Longitudinal changes in stimulation assays were assessed using linear mixed-effects models, with planned contrasts of day 90 and day 180 vs. day 0. P values were FDR-adjusted; only adjusted p < 0.05 are indicated. Data were derived from flow cytometry.

**Supplementary Figure 15.**
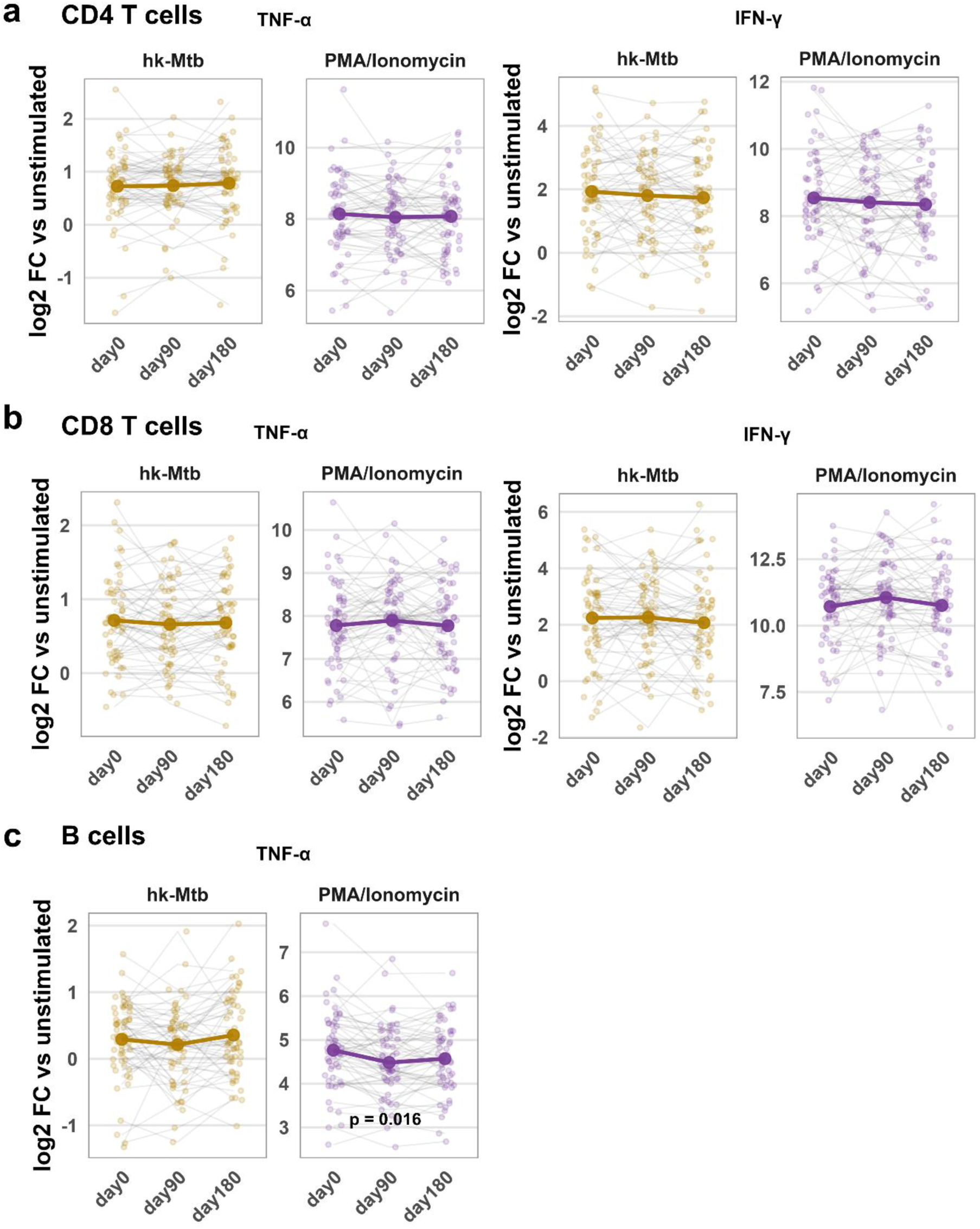
Cytokine production by lymphocyte subsets following stimulation. a) Log2 fold-changes relative to the corresponding unstimulated condition of TNF-α- and IFN-γ-producing CD4^+^ T cells following 4 h brefeldin A treatment under heat-killed *M. tuberculosis* (hk-Mtb) and PMA/Ionomycin stimulation. Thick line represents estimated marginal means and points with connecting thin lines represent individual participants (n = 55). b) Log2 fold-changes relative to the corresponding unstimulated condition of TNF-α- and IFN-γ-producing CD8^+^ T cells following 4 h brefeldin A treatment under hk-Mtb and PMA/Ionomycin stimulation. Thick line represents estimated marginal means and points with connecting thin lines represent individual participants (n = 55). c) Log2 fold-changes relative to the corresponding unstimulated condition of TNF-α-producing B cells following 4 h brefeldin A treatment under hk-Mtb and PMA/Ionomycin stimulation. Thick line represents estimated marginal means and points with connecting thin lines represent individual participants (n = 55). Statistics: Longitudinal changes in stimulation assays were assessed using linear mixed-effects models, with planned contrasts of day 90 and day 180 vs. day 0. P values were FDR-adjusted; only adjusted p < 0.05 are indicated. Data were derived from flow cytometry.

**Supplementary Figure 16.**
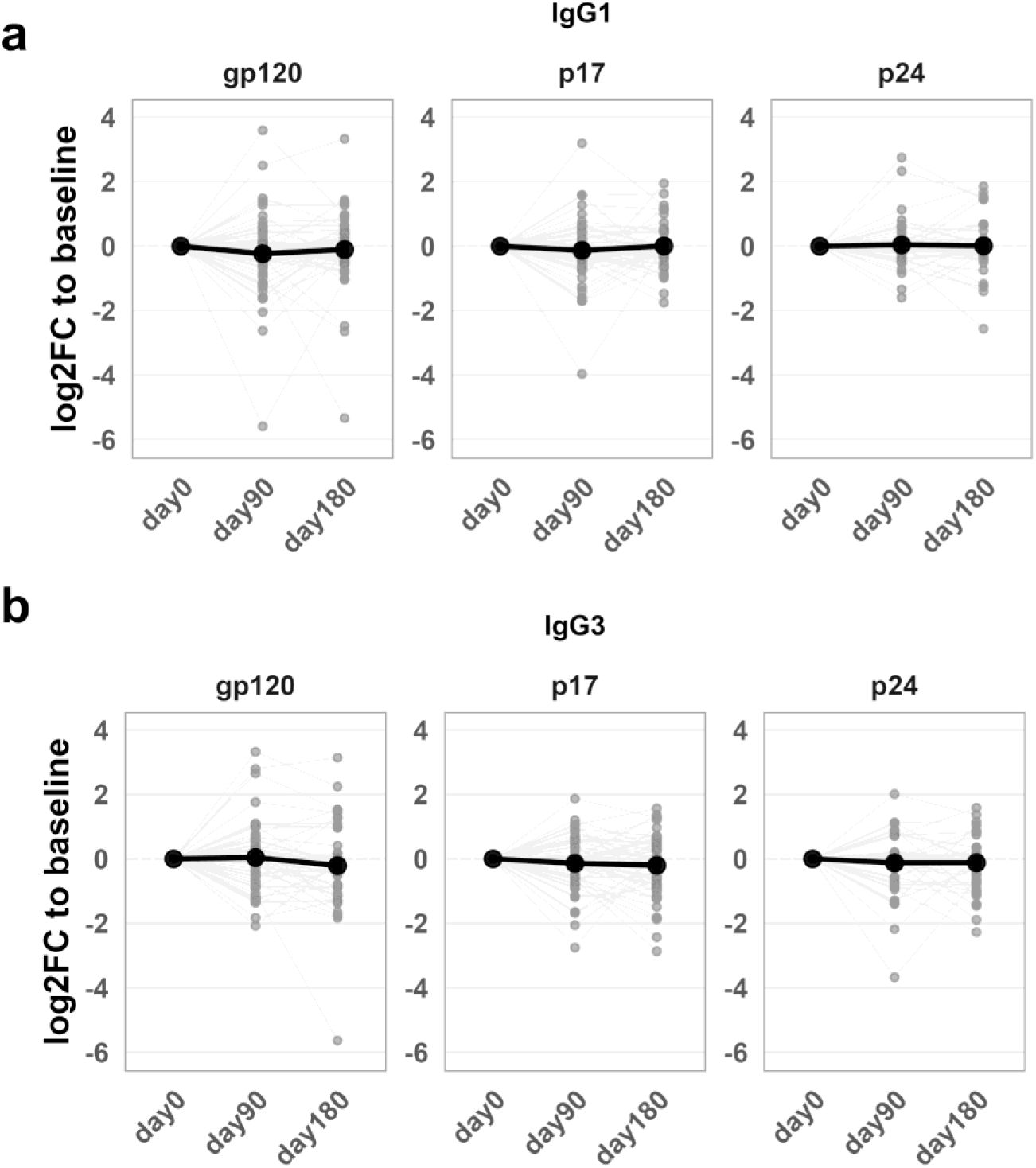
No evidence of systematic change in HIV-specific binding antibody responses over time. Log2 fold-changes relative to baseline (day 0) of HIV-specific binding antibody responses measured by binding antibody multiplex assay (BAMA) for gp120, p17, and p24, shown separately for IgG1 (a) and IgG3 (b). Thick line represents estimated marginal means and points with connecting thin lines represent individual participants (n = 55). Statistics: Longitudinal changes were assessed using linear mixed-effects models, with planned contrasts of day 90 and day 180 vs. day 0. P values were FDR-adjusted; only adjusted p < 0.05 are indicated. Data were derived from plasma binding antibody measurements.

**Supplementary Figure 17.**
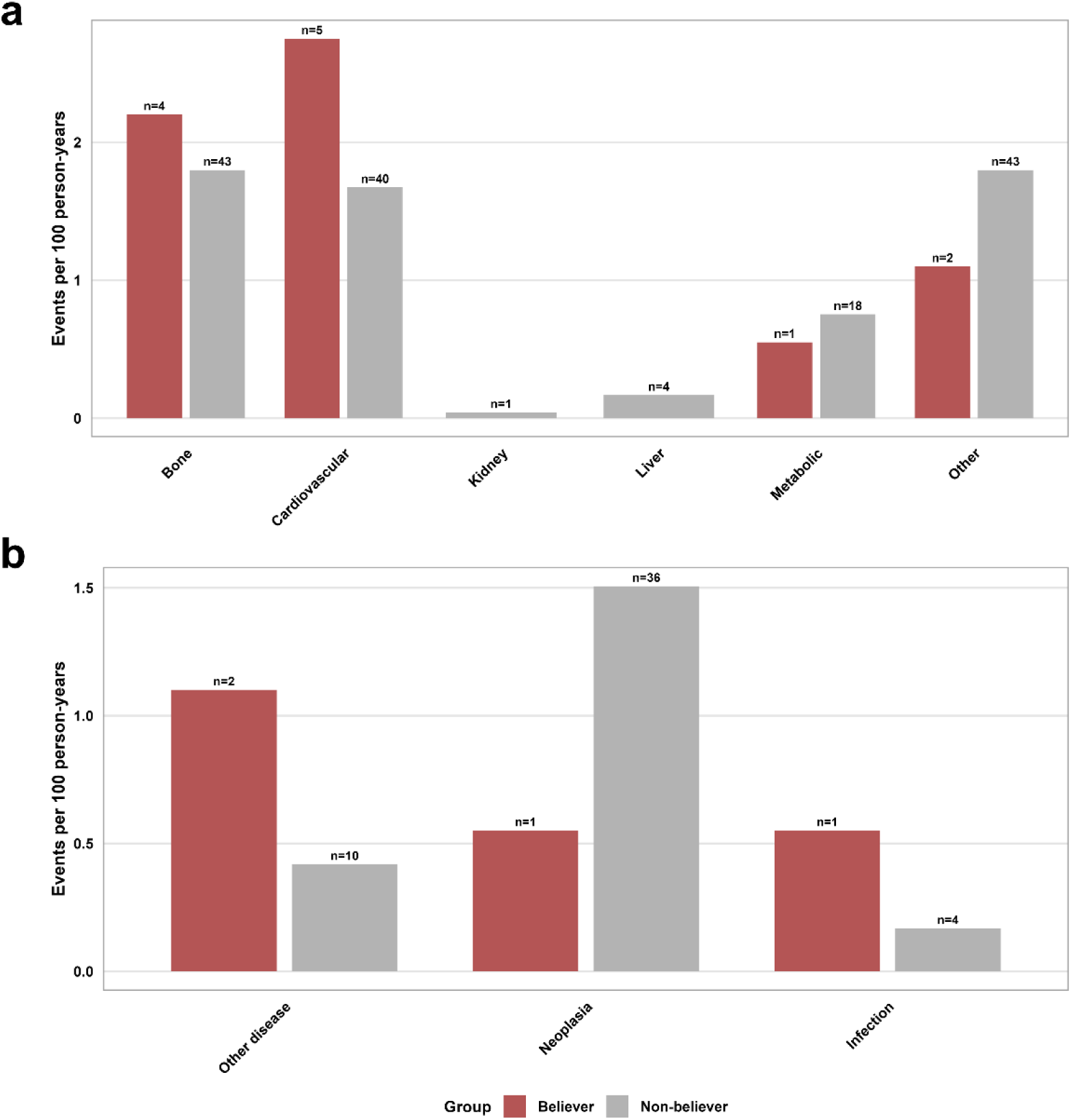
Clinical and disease-event rates in study participants and non-participants. a) Clinical event rates after BCG vaccination or the corresponding reference date, grouped by clinical category and normalized to 100 person-years. b) Disease event rates after BCG vaccination or the corresponding reference date, grouped as other disease, neoplasia, or infection-related events and normalized to 100 person-years. Statistics: Participant-level clinical and disease event rates were compared between Believer (n = 55) and Non-believer (n = 751) using Wilcoxon rank-sum tests. P values were FDR-adjusted; only adjusted p < 0.05 are indicated. These analyses should be interpreted descriptively given the low number of events. Data were derived from routine clinical records.

**Supplementary Figure 18.**
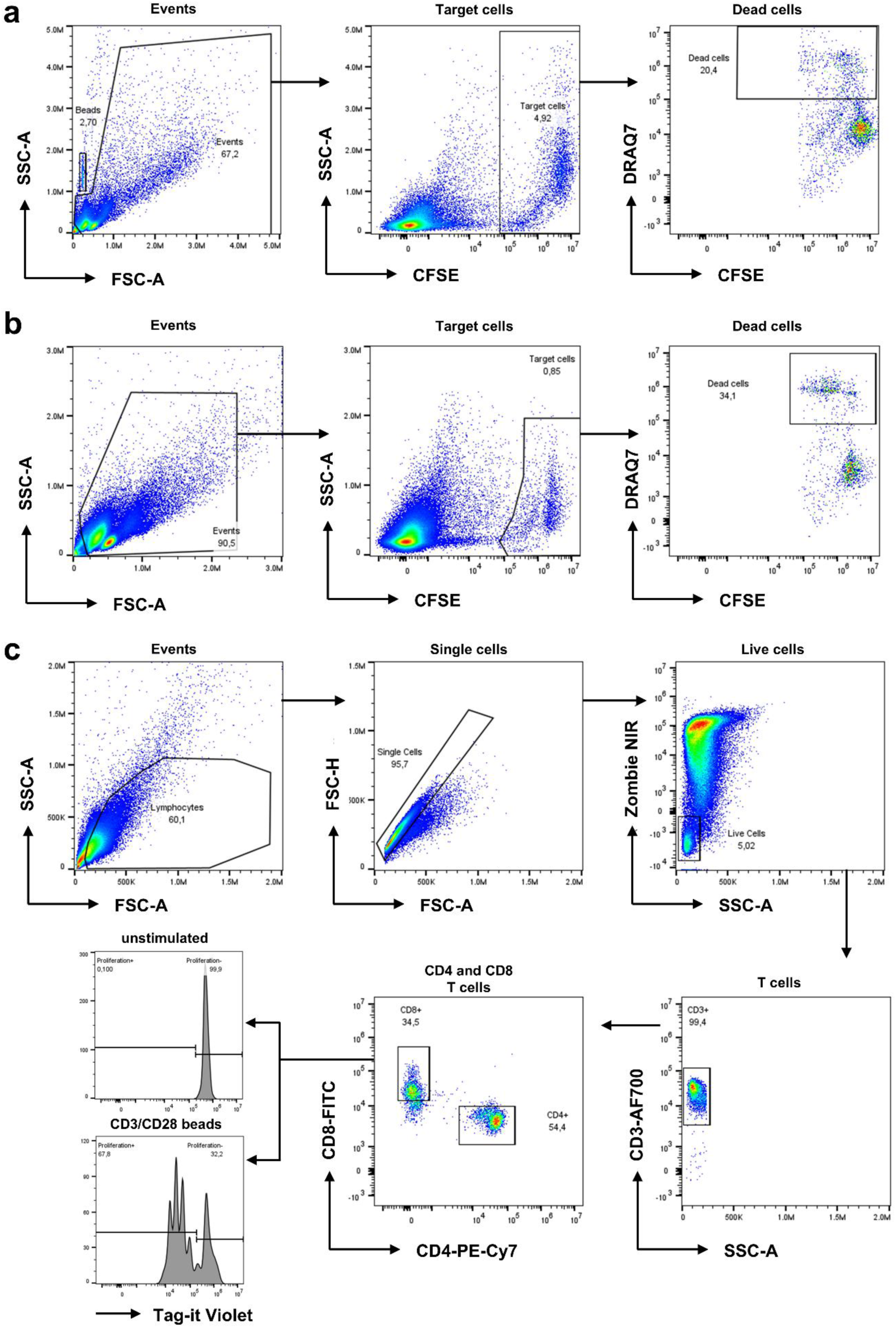
Flow cytometry gating strategies of in vitro assays. a) Representative flow cytometry plots of the cytotoxicity assay using K562 target cells at day 0. b) Representative flow cytometry plots of the antibody-dependent cellular cytotoxicity (ADCC) assay using Raji target cells in the rituximab-negative condition at day 0. c) Representative flow cytometry plots of the T cell proliferation assay in the unstimulated condition at day 0. The final histograms show Tag-it Violet fluorescence in unstimulated cells (top) and CD3/CD28 bead-stimulated cells (bottom).

**Supplementary Table 1.** Participant characteristics and HIV RNA burden by group.

| Marker | Fluorophore | Dilution factor |
| --- | --- | --- |
| <u>Extracellular</u> |  |  |
| CD11c | PE-CF594 | 50 |
| CD127 | PE-Cy7 | 400 |
| CD14 | StarBright UltraViolet 575 | 50 |
| CD16 | Brilliant UltraViolet 396 | 100 |
| CD19 | Brilliant Violet 650 | 100 |
| CD25 | PE-Fire 700 | 50 |
| CD3 | Brilliant Violet 570 | 50 |
| CD33 | StarBright UltraViolet 795 | 50 |
| CD38 | StarBright UltraViolet 740 | 50 |
| CD4 | APC-Fire 810 | 200 |
| CD40 | Brilliant Blue 515 | 200 |
| CD56 | Super Bright 780 | 100 |
| CD62L | StarBright Violet 515 | 50 |
| CD8a | Brilliant UltraViolet 615 | 50 |
| HLA-DR | APC-R700 | 200 |
| PD-1 | APC | 200 |
| Tim-3 | Brilliant Violet 421 | 50 |
| <u>Intracellular</u> |  |  |
| Granzyme B | RealBlue 780 | 100 |
| IFN-γ | eFluor 450 | 25 |
| TNF-α | PE | 100 |

